# Clonal Hematopoiesis Accelerates Frailty and Functional Decline

**DOI:** 10.64898/2026.09.24.26363888

**Authors:** Kun Zhao, Candice Quin, Yash Pershad, Liying Xue, Weizhi Cao, Grace I Bowman, Caitlyn Vlasschaert, Robert W Corty, J. Brett Heimlich, Erica N DeJong, Caitlin Lee, Zil Patel, Marco M. Buttigieg, Chris P Verschoor, Calum A Bane, Amy J.M. McNaughton, Dawn M.E. Bowdish, George A Kuchel, Eric A Whitsel, Aladdin H Shadyab, Charles Kooperberg, Andrea Z LaCroix, Alexander P Reiner, Michael J Rauh, Luke M Evans, Alexander G Bick

## Abstract

Frailty reflects declining physiological reserve across multiple organ systems with aging, yet the underlying biological mechanisms contributing to frailty remain poorly understood. We tested whether somatic mutations in the blood were associated with incident frailty and faster progression in 730,088 participants from UK Biobank, All of Us, BioVU and the Women’s Health Initiative. We found that clonal hematopoiesis of indeterminate potential (CHIP) and autosomal mosaic chromosomal alterations showed strong associations with frailty, whereas mosaic loss of chromosome X or Y did not. Findings were consistent across survey-based, directly measured and clinically assessed frailty, as well as healthspan phenotypes. A *Tet2* mouse model demonstrated multisystem frailty-like functional decline and T-cell remodeling. In humans, T-cell activation-related proteins directly mediated the association between clonal hematopoiesis and frailty. Collectively, these findings support a model in which clonal hematopoiesis accelerates biological aging through immunosenescence.

---

With age some individuals maintain physiologic resilience while others develop functional decline.^1–5^ Frail individuals have reduced reserve across multiple organ systems and are at high risk for disability, hospitalization, and death.^6–9^ Frailty can be quantified in multiple ways, including electronic health record–based scores^10^, self-reported deficit indices^11^, directly measured physical performance phenotypes^12^, and clinical assessments^13^. Despite advances in identifying frail individuals, the underlying biological processes that drive frailty in humans remain largely unknown.^7^ Prior studies have implicated chronic low-grade inflammation, immune dysregulation, sarcopenia, and impaired tissue repair as cellular characteristics of frailty.^14–16^ However, the upstream factors that initiate or amplify these processes in some individuals but not others are poorly defined.

Age-related clonal hematopoiesis (CH) represents a plausible upstream contributor linking aging biology to frailty. With age, dividing hematopoietic stem cells acquires an increasing burden of mutations. A subset of these mutations can confer a proliferative advantage giving rise to CH.^17–19^ CH prevalence increases sharply with age, affecting more than 10% of adults over 60.^20^ Although CH was initially recognized as a pre-cancer and linked to hematologic malignancy, subsequent studies have identified associations with a range of chronic, aging-related diseases and all-cause mortality.^21–25^ Multiple classes of mutations drive CH, including mutations in myeloid cancer driver genes that give rise to Clonal Hematopoiesis of Indeterminate Potential (CHIP), large-scale chromosomal mosaicism termed mosaic Chromosomal Abnormalities (mCAs), and loss of sex chromosomes X and Y.

Given that CH has been associated with altered immune-cell function and inflammatory signaling processes which have been posited to contribute to frailty, we hypothesized that CH may increase risk of incident frailty and its progression.^26,27^ To test this hypothesis, we analyzed 726,091 participants across three large biobanks with linked genomic and healthcare data—the UK Biobank (UKB), the All of Us (AoU) Research Program, and Vanderbilt BioVU. We evaluated the associations of CH with incident frailty and longitudinal frailty progression. We further assessed the consistency of these associations across five complementary measures of frailty and healthy aging^11–13,28^, including a standardized measure of physical frailty in the Women’s Health Initiative (WHI) and clinically assessed functional impairment in three human aging cohorts. To further investigate potential mechanisms linking CH to frailty, we leveraged an *in vivo* murine model of clonal hematopoiesis and plasma proteomic data from UKB (**Extended Data Fig.1**).

## Results

After applying the inclusion and exclusion criteria, 726,091 participants aged 40-70 years were included across three cohorts: 130,039 from AoU, 111,523 from BioVU, and 484,529 from UKB (**Table 1**, **Figure 1A**). Median age at blood sampling ranged from 55 to 58 years, and women comprised 54%-62% of participants. Most participants were of European ancestry, particularly in UKB (95.3%), whereas AoU and BioVU included larger proportions of participants of African ancestry (21.5% and 11.8%, respectively). Frailty was quantified in these cohorts using the Hospital Frailty Risk Score (HFRS)^10^, a deficit accumulation Frailty Index which was originally developed in individuals aged 75+ and subsequently validated across all adult age groups^29^.

**Figure 1.**
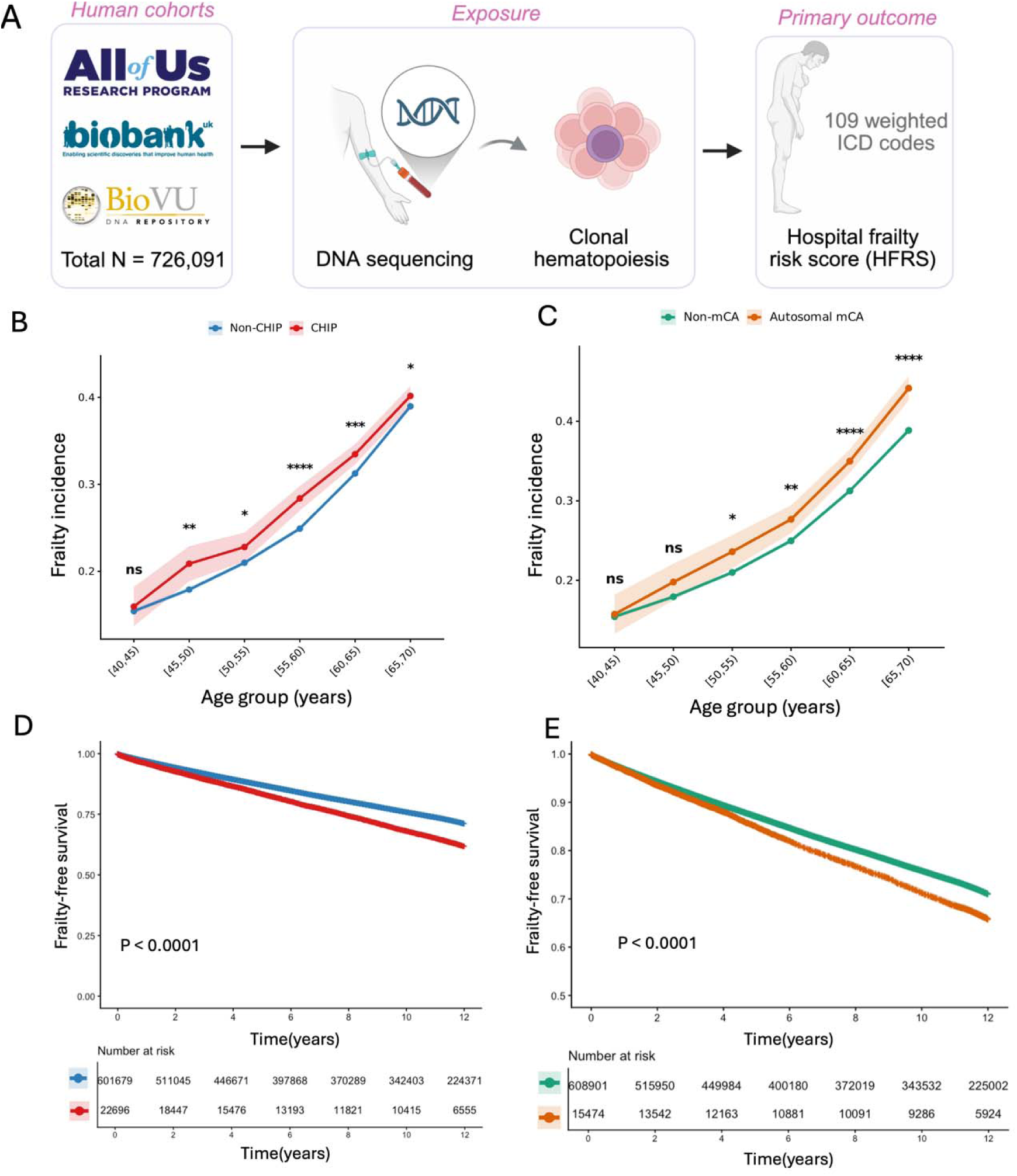
Age-specific frailty incidence and frailty-free survival according to clonal hematopoiesis status. (**A**) Primary study design across three large human biobanks, including the All of Us Research Program, UK Biobank, and Vanderbilt BioVU. Age-specific frailty incidence stratified by **(B)** CHIP and **(C)** autosomal mCA status. Frailty incidence increased progressively with age in both groups, with consistently higher incidence observed among individuals with CHIP across age strata. Kaplan–Meier curves for frailty-free survival according to **(D)** CHIP and **(E)** autosomal mCA status. Numbers at risk are shown below each panel.

**Table 1.** Baseline Characteristics of Participants Across Three Large Human Cohorts.

|  | NIH All of Us | Vanderbilt BioVU | UK BioBank |
| --- | --- | --- | --- |
| N | 130,039 | 111,523 | 484,529 |
| Median age at blood sample (IQR) | 57 (50-64) | 55 (48-62) | 58 (50-63) |
| N women (%) | 81,086 (62.4%) | 60,686 (54.4%) | 262,001 (54.1%) |
| Ever smoked (%) | 54,926 (42.2%) | 40,317 (36.2%) | 218,182 (45%) |
| Ancestry |  |  |  |
| *European* | 76,304 (58.7%) | 92,464 (82.9%) | 461,711 (95.3%) |
| *African* | 28,018 (21.5%) | 13,169 (11.8%) | 9,255 (1.9%) |
| *East Asian* | 2,114 (1.6%) | 1,154 (1.0%) | 13,563 (2.8%) |
| N with clonal hematopoiesis of indeterminate potential (%) |  |  |  |
| *Overall CHIP* | 4,996 (3.8%) | 6,331 (5.7%) | 15,702 (3.2%) |
| *DNMT3A* | 2,820 (2.2%) | 3,281 (2.9%) | 9,630 (2.0%) |
| *TET2* | 843 (0.6%) | 1,517 (1.4%) | 2,334 (0.5%) |
| *ASXL1* | 331 (0.3%) | 616 (0.6%) | 1,823 (0.4%) |
| *JAK2* | 90 (0.07%) | 104 (0.09%) | 159 (0.03%) |
| N with mosaic chromosomal alterations (%) |  |  |  |
| *Autosomal mCA* | 2,636 (2.03%) | 631 (0.57%) | 14,807 (3.06%) |
| *mLOX* | 2,887 (2.22%) | 68 (0.06%) | 14,879 (3.32%) |
| *mLOY* | / | 2,390 (2.14%) | 41,783 (9.32%) |
| Hospital Frailty Risk Score (HFRS) | 3.4 (0.5-7.6) | 2.0 (0-6.3) | 3.1 (0-7.8) |
| N with frailty (>5) | 49,909 (38.4%) | 34,370 (30.8%) | 180,559 (37.3%) |
| N with severe frailty (>15) | 10,783 (8.3%) | 8,200 (7.4%) | 46,415 (9.6%) |
| Median years of follow-up (IQR) | 2.8 (1.1-4.1) | 4.9 (2-11.1) | 12.4 (11.7-13.1) |

Data are presented as median (interquartile range [IQR]) for continuous variables and number (percentage) for categorical variables. Frailty risk score was calculated using the Hospital Frailty Risk Score (HFRS) algorithm. Frailty was defined as HFRS > 5 and severe frailty as HFRS > 15. Mosaic chromosomal alterations (mCA) were classified as autosomal mCA, mosaic loss of chromosome X (mLOX), and mosaic loss of chromosome Y (mLOY). mLOY was not available in the All of Us cohort. Follow-up time was calculated from the date of blood sampling to the last available clinical record.

Median HFRS ranged from 2.0 to 3.4, 31%-38% of participants met the predefined criterion for frailty^10,30^ (HFRS >5), and 7%-10% met criteria for severe frailty (HFRS >15). These frequencies were broadly consistent with previous large population-based studies.^29,31^

Any clonal hematopoiesis, defined by the presence of CHIP and/or an mCA, was identified in 5.7% of participants in AoU, 6.1% in BioVU, and 6.2% in UK Biobank. CHIP prevalence ranged from 3.2% to 5.7% across cohorts, with *DNMT3A* being the most common driver gene, followed by *TET2* and *ASXL1*; *JAK2* mutations were rare (<0.1% in all cohorts). Among mCA subtypes, autosomal mCAs were detected in 0.6%-3.1% of participants across cohorts. The prevalence of both CHIP and mCAs increased progressively with age (**Extended Data Fig.2**).

**Figure 2.**
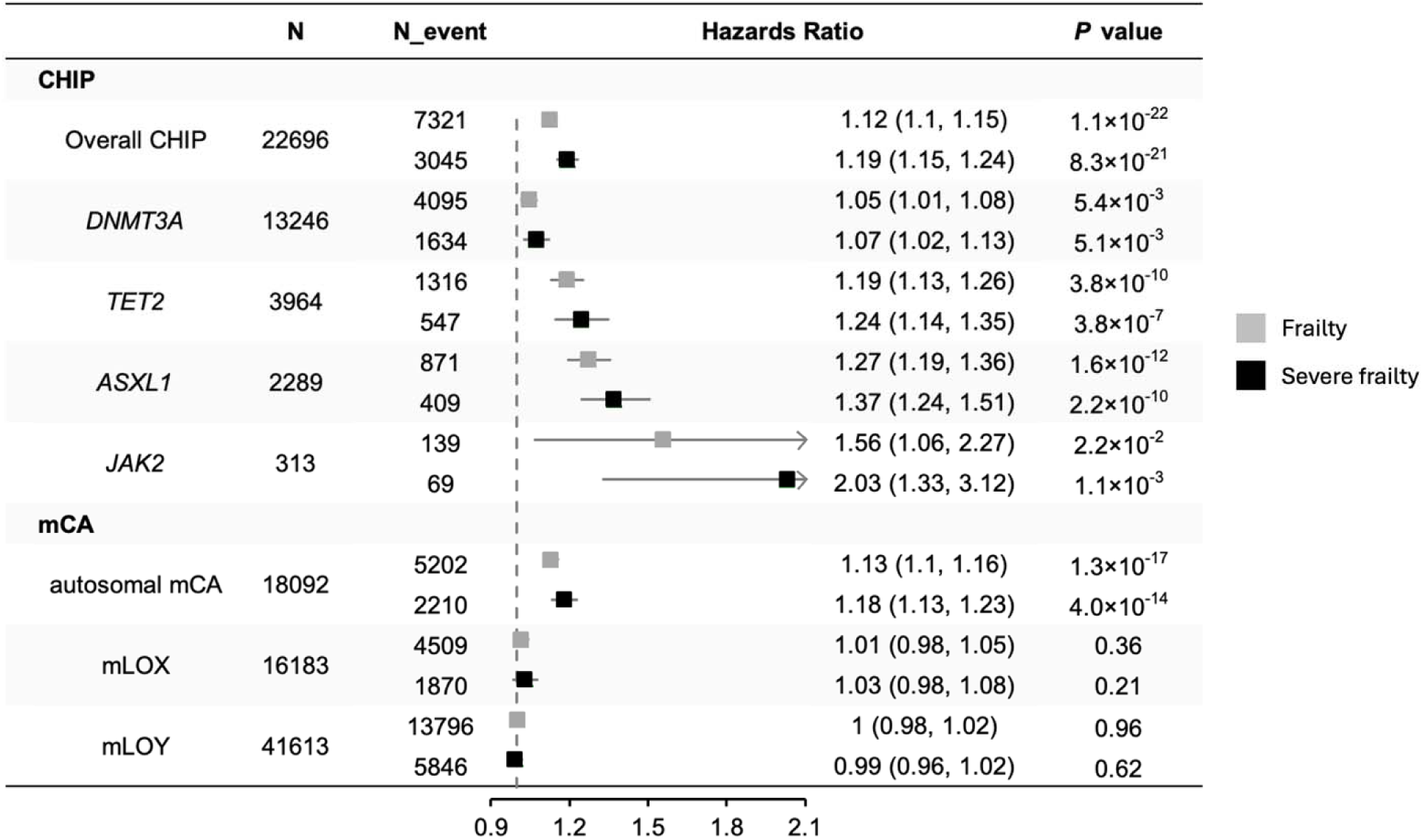
Associations of clonal hematopoiesis with incident frailty. Forest plot showing hazard ratios (HRs) and 95% confidence intervals for incident frailty (grey squares) and severe frailty (black squares) according to CHIP status, gene-specific CHIP subtypes, and mCA (autosomal mCA, mLOX and mLOY). Estimates represent meta-analyses across UK Biobank, All of Us, and BioVU cohorts. Within each cohort, Cox proportional hazards models were adjusted for age, age^2^, sex, smoking status, and genetic principal components (PC1-PC5). Numbers of carriers (N), Numbers of events among carriers (N_event), hazard ratios with 95% confidence intervals, and corresponding *P* values are shown.

### Age-specific frailty incidence and frailty-free survival by clonal hematopoiesis status

Across all age groups, individuals with CHIP exhibited a higher incidence of frailty than non-carriers, with statistically significant differences from age 45 onward (**Figure 1B**). The absolute difference between CHIP and non-CHIP participants was greatest between ages 55 and 60.

Similarly, autosomal mCAs were associated with higher frailty incidence, with statistically significant differences observed from age 50 onward and increasing separation after age 55 (**Figure 1C**). In contrast, the age-specific incidence curves for mLOY and mLOX, in males and females respectively, largely overlapped with those of their reference groups at younger ages, with significant differences becoming apparent only in older age groups (**Extended Data Fig. 3**).

Kaplan–Meier analyses demonstrated significantly lower frailty-free survival among individuals with CHIP (**Figure 1D**) and autosomal mCA (**Figure 1E**) compared with their respective reference groups (log-rank *P* < 0.0001). At 10 years, the estimated cumulative incidence of frailty was 31.8% among CHIP carriers compared with 24.0% among non-CHIP carriers.

Similarly, the 10-year cumulative incidence was 28.7% among autosomal mCA carriers compared with 24.1% among non-carriers. Unadjusted frailty-free survival curves for mLOY and mLOX also differed statistically from their reference groups (log-rank *P* < 0.0001); however, the absolute differences were modest, particularly for mLOX, with 10-year cumulative frailty incidences of 26.6% versus 24.5% (**Extended Data Fig.4**).

### CHIP is associated with increased frailty risk in a gene- and clone-dependent manner

In multivariable-adjusted time-to-event analyses, CHIP was consistently associated with a higher risk of incident frailty across AoU, BioVU, and UKB (**Supplementary Table 4)**. In fixed-effects meta-analysis, overall CHIP was associated with a 12% higher risk of incident frailty (HR 1.12, 95% CI 1.10-1.15, I^2^ = 4.6%, *P* = 1.1×10^-22^) and a 19% higher risk of severe frailty (HR 1.19, 95% CI 1.15-1.24, I^2^ = 17.6%, *P* = 8.3×10^-21^) (**Figure 2**; **Supplementary Table 5**). Driver gene– specific analyses demonstrated substantial heterogeneity in frailty risk across CHIP subtypes (*P* for heterogeneity = 1.1 × 10^-8^). *DNMT3A* mutations were associated with modest increases in risk for both incident frailty (HR 1.05, 95% CI 1.01-1.08) and severe frailty (HR 1.07, 95% CI 1.02–1.13). In contrast, *TET2* and *ASXL1* mutations had larger point estimates. *TET2* mutations were associated with a 19% higher risk of incident frailty (HR 1.19, 95% CI 1.13-1.26) and a 24% higher risk of severe frailty (HR 1.24, 95% CI 1.14-1.35). Similarly, *ASXL1* mutations were associated with 27% and 37% higher risk of frailty and severe frailty, respectively (HR 1.27, 95% CI 1.19-1.36; HR 1.37, 95% CI 1.24-1.51). *JAK2* mutations exhibited the largest effect size, particularly for severe frailty (HR 2.03, 95% CI 1.33-3.12), although estimates were based on relatively small numbers (n = 69). Notably, across all driver genes, the effect sizes were consistently higher for severe frailty than for overall frailty.

We next examined whether clone size, quantified by variant allele fraction (VAF), was associated with frailty risk. When analyses were restricted to large clones (variant allele fraction [VAF] >10%), the overall pattern remained consistent, with generally stronger effect estimates observed (**Supplementary Table 4-5)**. However, the association for *DNMT3A*-CHIP was no longer statistically significant (**Extended Data Fig.5**). Findings remained consistent in sensitivity analyses additionally adjusting for baseline HFRS (**Supplementary Table 6**).

### mCAs of the autosomes, but not sex chromosomes, are associated with increased frailty risk

Autosomal mCAs were associated with an increased covariate-adjusted risk of incident frailty (HR 1.13, 95% CI 1.10–1.16), with a larger effect estimate observed for severe frailty (HR 1.18, 95% CI 1.13–1.23) (**Figure 3**). Although unadjusted Kaplan–Meier analyses showed modest differences in frailty-free survival for both mLOX in females and mLOY in males, neither of them was significantly associated with frailty incidence in adjusted analyses (*P* > 0.21). We further evaluated autosomal mCAs at the chromosome arm-specific level. Five recurrent autosomal mCA events were significantly associated with increased frailty incidence, including chr14 CN-LOH, chr11q Loss, chr9p CN-LOH, chr4q CN-LOH, and chr13q Loss (HR 1.39-1.73; *P* < 1.03×10^-4^) (**Extended Data Fig.6; Supplementary Table 7**). These events spanned both lineage-associated mCA classes: chr11q and chr13q losses are characteristic lymphoid-associated alterations commonly observed in chronic lymphocytic leukemia, whereas chr9p and chr4q CN-LOH are myeloid-associated events involving the *JAK2* and *TET2* loci, respectively; chr14 CN-LOH has also been classified as a myeloid-associated mCA^32^.

**Figure 3.**
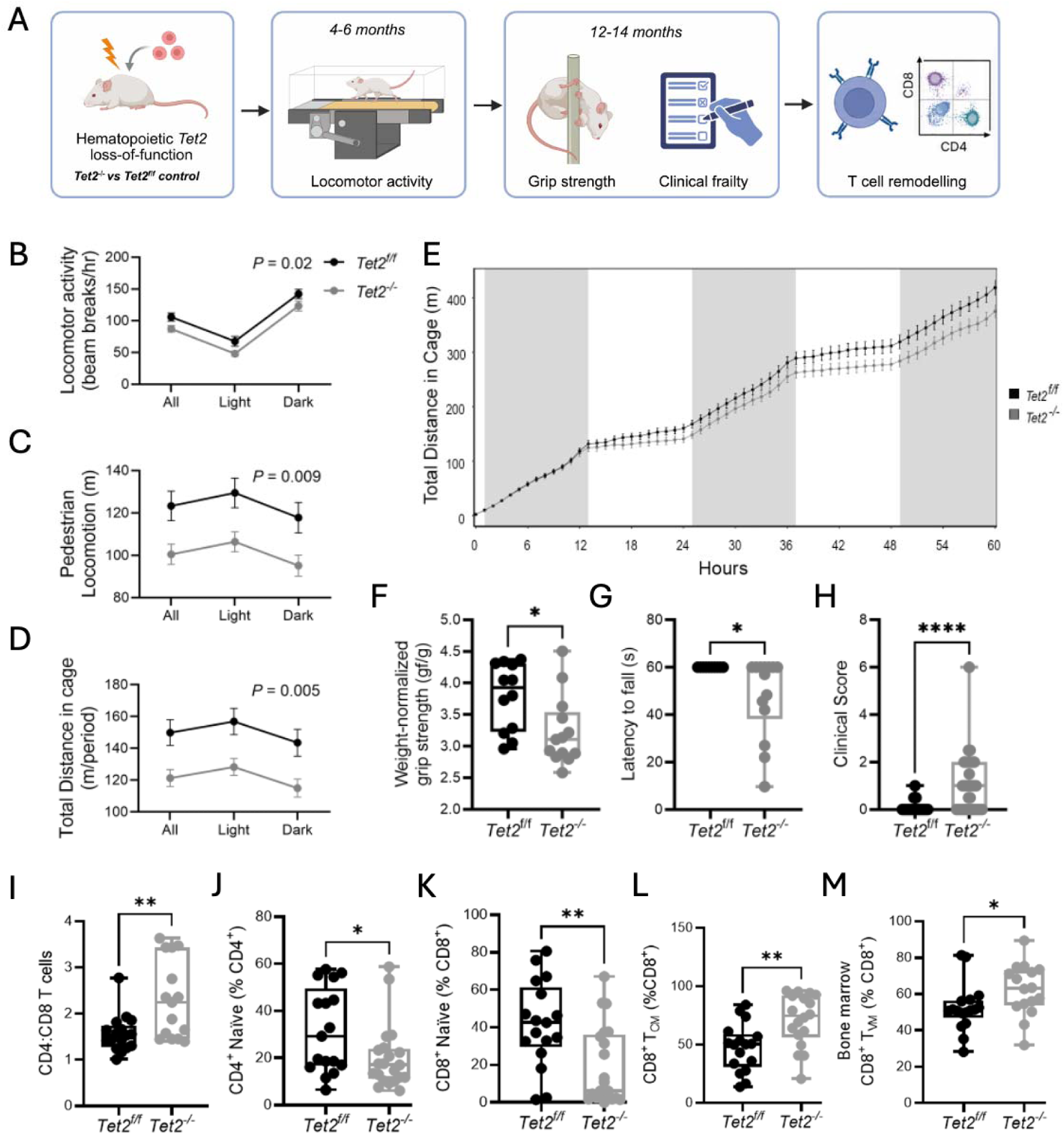
Hematopoietic Tet2 loss-of-function is associated with reduced locomotor activity and frailty-like functional decline in mice. **(A)** Experimental overview. **(B–E)** At 4–6 months, *Tet2^-/-^*mice showed reduced spontaneous locomotor activity and cumulative distance travelled compared with *Tet2^f/f^* controls (*Tet2^f/f^*, n = 19; *Tet2^-/-^*, n = 21). **(F–H)** At 12–14 months, *Tet2^-/-^* mice exhibited reduced weight-normalized grip strength, impaired cage-hang performance, and higher clinical frailty scores. **(I–M)** Flow cytometry demonstrated an increased CD4:CD8 ratio, reduced naïve CD4^+^ and CD8^+^ T-cell proportions, and increased CD8^+^ central-memory and bone marrow virtual-memory T-cell proportions. *P* values are shown in the figure; \**P* ≤ 0.05, \*\**P* ≤ 0.01, \*\*\**P* ≤ 0.0001.

### Mutation-specific risk of incident frailty

Subtype-specific analyses demonstrated marked heterogeneity in effect estimates across both CHIP driver genes and autosomal mCAs (**Extended Data Fig.6**). For *DNMT3A*-CHIP, overall associations appeared to be driven primarily by missense mutations (n = 7,306) rather than loss-of-function variants (n = 4,553). *DNMT3A* missense mutations were associated with increased frailty risk (HR 1.10, 95% CI 1.03-1.17; *P_FDR_* = 7.32×10^-3^), whereas loss-of-function variants did not reach statistical significance (HR 1.19, 95% CI 1.00-1.41; *P_FDR_* = 0.07). Further stratification of missense mutations demonstrated that the association was largely attributable to the R882* hotspot (n = 1,575), which showed a stronger effect (HR 1.32, 95% CI 1.16-1.49; *P_FDR_* = 8.36×10^-5^), while non-R882 missense variants were not significantly associated with frailty (HR 1.04, 95% CI 0.96-1.12; *P_FDR_* = 0.33). In contrast, for *TET2*-CHIP, the signal was primarily driven by loss-of-function mutations (HR 1.38, 95% CI 1.23-1.54; *P_FDR_* = 2.84×10^-8^, n = 2,345), whereas missense variants were not significantly associated with incident frailty (HR 1.15, 95% CI 0.97-1.35; *P_FDR_*= 0.11, n = 1,081).

### CHIP and autosomal mCAs are associated with accelerated frailty progression

Given the association between CHIP and autosomal mCAs and incident frailty, we next investigated how they alter change of frailty trajectory over time. Both CHIP and autosomal mCAs were associated with accelerated frailty trajectories in longitudinal analyses. Overall CHIP was associated with a faster increase in HFRS over time, corresponding to an additional 0.046 HFRS units per year (β = 0.046, SE = 0.016; *P* = 5.45×10^-3^) (**Extended Data Fig.7**). Similar associations were observed for *DNMT3A*-CHIP (β = 0.037, SE = 0.013; *P* = 4.41×10^-3^), *TET2-*CHIP (β = 0.061, SE = 0.024; *P* = 1.26×10^-2^) and *ASXL1-*CHIP (β = 0.076, SE = 0.028; *P* = 6.73×10^-3^). *JAK2*-CHIP showed the largest effect estimate (β = 0.101, SE = 0.051; *P* = 4.86×10^-2^), although precision was limited by smaller carrier numbers. The relative ranking of CHIP driver genes was preserved across longitudinal trajectory and incident frailty analyses. Similarly, autosomal mCAs were associated with greater longitudinal frailty progression (β = 0.037, SE = 0.007; *P* = 3.58×10^-7^). In contrast, neither mLOX nor mLOY was significantly associated with frailty trajectory (*P*≥0.20).

### CHIP associations extend across diverse measures of frailty and healthspan

To determine whether these findings were robust across complementary measures of functional aging, we evaluated CHIP associations using three additional phenotypes in the UK Biobank. Using a recently developed electronic frailty index (eFI)^28^, repeated-measures analyses showed that overall CHIP was associated with accelerated eFI progression (**Supplementary Table 8**). Similar associations were observed for *DNMT3A*-, *TET2*-, *ASXL1*-, and *JAK2*-CHIP (all *P* < 1.62×10^-5^). Autosomal mCAs also showed significant associations with faster eFI progression (*P* = 4.51×10^-60^).

We next examined a survey-based frailty index derived from 49 self-reported health deficits^11^. CHIP carriers had higher frailty index values than non-carriers (*P* < 0.001; **Extended Data Fig. 8**), and this association remained significant after adjustment for age and sex (β = 0.003, 95% CI 0.002-0.004; *P* < 0.001). We also evaluated the Healthspan Proteomic Score (HPS)^33^, which integrates chronological age and 86 circulating proteins to estimate the probability of remaining free from major healthspan-ending conditions. Among 48,874 participants with available data, CHIP carriers had lower scores than non-carriers (median, 0.74 versus 0.82; *P* < 0.001; **Extended Data Fig. 9**), indicating a lower predicted probability of maintaining healthspan. This association remained significant after adjustment for age and sex (β = -0.018, 95% CI -0.025 to - 0.012; *P* < 0.001). Together, these findings demonstrate that the association between CHIP and functional aging extends across electronic, survey-based, and proteomic measures of frailty and healthspan.

### Independent cohorts support associations between CHIP and clinically assessed frailty

We next examined the association between CHIP and frailty in independent cohorts using directly or clinically assessed frailty measures. In the 3,997 participants from Women’s Health Initiative Long Life Study (WHI-LLS)^34^ (**Supplementary Table 9)**, frailty was assessed using the Fried phenotype^12^. CHIP status measured at baseline showed a directionally consistent association with incident frailty at first LLS visit (LLS-1) (OR 1.17, 95% CI 0.86–1.60; *P* = 0.32). A similar pattern was observed for *TET2*-CHIP (OR 1.22, 95% CI 0.85–1.74; *P* = 0.29), whereas *DNMT3A*-CHIP was not associated with incident frailty (OR 1.03, 95% CI 0.55–1.93; *P* = 0.92). These odds ratios were consistent with the HFRS risk estimates.

To complement the prospective analysis, we next performed cross-sectional analyses relating CHIP to prevalent Fried frailty measured at LLS-1. CHIP defined at VAF ≥2% was not significantly associated with prevalent frailty (*P*>0.05). However, when analyses were restricted to larger clones with VAF >10%, overall CHIP was associated with higher odds of frailty (OR 1.41, 95% CI 1.01–1.96; *P* = 0.044). *TET2*-CHIP was similarly associated with frailty (OR 1.71, 95% CI 1.12–2.61; *P* = 0.012), whereas *DNMT3A*-CHIP was not (OR 0.99, 95% CI 0.56–1.75) (**Extended Data Fig.10**). The limited number of incident frailty cases, attrition between visits, and potential survivor bias in the approximately 15 years between baseline and LLS-1 may have contributed to the downwardly biased association estimates.

We further evaluated this association in three human aging cohorts^35^, comprising 205 adults aged 65 years or older with available Clinical Frailty Scale assessments (**Supplementary Table 10**). The proportion of CHIP carriers increased progressively across higher CFS categories (*P* = 0.04), with the highest prevalence observed among individuals with greater frailty severity (**Extended Data Fig.11**). Using cumulative link mixed model, CHIP was associated with higher odds of being in a greater Clinical Frailty Scale category (OR 2.05, 95% CI 1.02–4.13; *P* = 0.044).

Together, these analyses provide supportive evidence of the robustness of the association between CHIP and frailty, given that it extends across directly measured physical frailty and clinically assessed frailty phenotypes in multiple independent cohorts of older adults.

### Hematopoietic Tet2 loss-of-function accelerates frailty-like decline and immune aging in mice

To directly test whether CHIP drives frailty, we modeled *Tet2-*mutant CHIP in mice. We examined mice with hematopoietic-specific *Tet2* loss-of-function (*Vav1-iCre^+^; Tet2^−/−^*) and *Tet2^f/f^* littermate controls (**Figure 3A**). At 4-6 months of age, *Tet2^−/−^*mice exhibited reduced spontaneous locomotor activity, including fewer locomotor beam breaks (*P* = 0.02), reduced ambulatory locomotion (*P* = 0.009), and shorter total distance travelled (*P* = 0.005) across the light and dark phases (*Tet2^f/f^,* n = 19; *Tet2^−/−^*, n = 21; **Figure 3B-D**). Consistently, cumulative distance travelled over the 60-hour recording period was lower in *Tet2^−/−^*mice than in controls (*P* = 0.023, **Figure 3E**). These differences were not accompanied by consistent changes in body mass, oxygen consumption, carbon dioxide production, or respiratory exchange ratio, suggesting that reduced activity was not solely attributable to differences in body size or basal metabolism. By middle age (12–14 months), *Tet2^−/−^* mice had lower weight-normalized grip strength than controls (3.2 ± 0.15 versus 3.7 ± 0.15 g/g; *P* ≤ 0.05) and shorter cage-hang duration (47.3 ± 4.8 versus 59.7 ± 0.27 seconds; *P* ≤ 0.05) (**Figure 3F-G**). They also had higher clinical frailty scores (0.92 ± 0.27 versus 0.058 ± 0.058; **Figure 3H**), based on a validated composite index assessing deficits across multiple physiological systems,^36^ consistent with accelerated multisystem functional decline.

Since *TET2* loss-of-function is known to bias hematopoietic differentiation away from lymphopoiesis^37^ and frailty is characterized by an immunosenescent phenotype marked by the accumulation of late-stage T cells,^16^ we reasoned that T-cell remodeling may be a plausible mechanism by which *TET2* CHIP exacerbated the frailty in mice. *Tet2^−/−^* mice showed an increased CD4:CD8 T-cell ratio, reduced proportions of naïve CD4^+^ and CD8^+^ T cells, and increased proportions of CD8^+^ central-memory T cells in peripheral blood and CD8^+^ virtual-memory T cells in bone marrow (**Figure 3I–M**). Additional analyses showed an increased monocyte-to-T-cell ratio, unchanged absolute naïve CD^+^ T-cell numbers, and reduced absolute numbers of naïve CD8^+^, CD8^+^ effector-memory, and CD183^+^CD8^+^ effector-memory T cells (**Extended Data Fig. 12**). Together, these findings indicate a shift from naïve toward differentiated memory T-cell states, consistent with diminished capacity to mount immune responses to novel pathogens and age-associated immune remodeling. By modelling *TET2*-mutant clonal hematopoiesis in mice, we demonstrate that hematopoietic Tet2 loss is sufficient to induce T-cell remodeling and frailty-like functional decline, providing experimental support for a causal contribution of *TET2*-mutant hematopoiesis to frailty.

### Immune remodeling links clonal hematopoiesis to frailty in humans

Having observed frailty-like functional decline accompanied by T-cell remodeling in mice, we next asked whether T-cell–related immune dysregulation represented a convergent mechanism linking clonal hematopoiesis to frailty in humans. To address this question, we analyzed Olink plasma proteomic data from UK Biobank, focusing on 148 proteins encoded by genes associated with T cell activation gene ontology (**Supplementary Table 11**). In Cox proportional hazards models adjusted for age, age squared, sex, smoking status, and genetic principal components 1–5, 100 of the 148 proteins were significantly associated with incident frailty after correction for multiple testing (**Figure 4A**). Most significant associations were positive, indicating that higher circulating levels of T cell activation–related proteins were associated with greater frailty risk.

**Figure 4.**
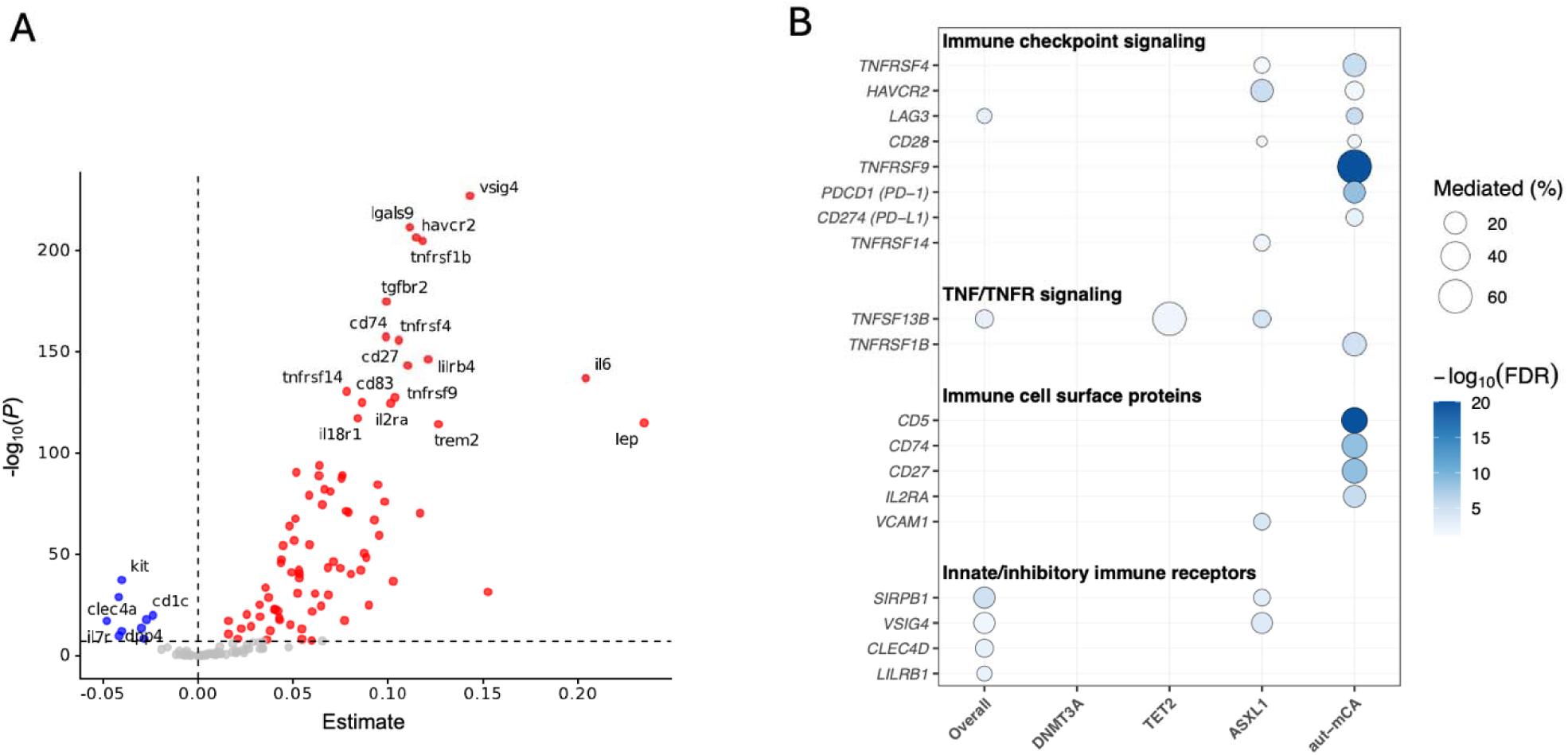
T-cell activation–related proteins are associated with frailty and mediate clonal hematopoiesis–frailty associations. **(A)** Volcano plot showing associations of 148 plasma proteins annotated to the Gene Ontology term T-cell activation with incident frailty in UK Biobank. Estimates were derived from Cox proportional hazards models adjusted for age, age squared, sex, smoking status, and genetic principal components 1–5. The horizontal dashed line indicates the multiple-testing significance threshold. Red points denote proteins positively associated with frailty, blue points denote inverse associations, and gray points denote nonsignificant associations. Selected proteins are labeled. **(B)** Bubble plot summarizing significant mediators of the associations between clonal hematopoiesis and incident frailty. Columns represent overall CHIP, *DNMT3A*-, *TET2*-, and *ASXL1*-CHIP, and autosomal mCAs (aut-mCA). *JAK2*-CHIP was not evaluated due to insufficient sample size. Proteins are grouped by functional category. Bubble size indicates the proportion of the total association mediated, and bubble color indicates -log10(FDR). Blank cells indicate no significant mediation after FDR correction.

Consistent with the increased CD4:CD8 T-cell ratio observed in mouse model, a higher Olink-derived CD4/CD8A protein ratio was associated with increased risk of incident frailty (HR 1.03, 95% CI 1.01–1.05; *P* = 3.4 × 10^-4^).

Among proteins associated with both CHIP and incident frailty, we performed mediation analyses to estimate the proportion of the CHIP–frailty association statistically explained by each protein (**Supplementary Table 12**). For overall CHIP, seven proteins remained significant after FDR correction, SIRPB1, VSIG4, TNFSF13B, CLEC4D, LILRB1, LAG3, and SLAMF1, with individual mediated proportions ranging from approximately 4% to 18%. Mediation patterns differed substantially across driver genes. No significant mediators were identified for *DNMT3A*-CHIP. TNFSF13B, the key driver of B lymphocyte activation (also known as BAFF), mediated 59.2% of the *TET2*-CHIP association (*P*_FDR_ = 0.014). *ASXL1*-CHIP was associated with 18 significant mediators, including several TNF/TNFR superfamily members. Autosomal mCAs also demonstrated substantial immune mediation, predominantly involving TNFR and CD family proteins; immune checkpoint receptor TNFRSF9, or CD137, showed the largest mediated proportion (63.1%), followed by CD5, CD74, and CD27 (**Figure 4B**).

## Discussion

In this study, we found that two forms of clonal hematopoiesis, CHIP and autosomal mCAs, were consistently associated with incident frailty and accelerated frailty progression. Overall CHIP was associated with a 12% higher risk of incident frailty and a 19% higher risk of severe frailty, whereas autosomal mCAs conferred similar risk estimates. Associations differed according to CHIP driver gene, mutation class, clone size, and mCA subtype. Importantly, these findings were robust across multiple complementary measures of functional aging. Finally, convergent evidence from a murine model and human plasma proteomics suggests that T-cell immunosenescence may represent one mechanism through which clonal hematopoiesis contributes to declining physiological reserve.

Several implications emerge from these findings. First, our results extend the clinical relevance of clonal hematopoiesis beyond individual age-related diseases and support its potential role as a contributor to frailty. CHIP has been linked to an expanding spectrum of age-related conditions, including hematologic malignancies^17,18,32,38^, cardiovascular disease^39–42^, chronic kidney disease^43,44^, chronic liver disease^45^ and osteoporosis^46^. In contrast to these individual disease outcomes, frailty reflects the cumulative loss of physiological reserve across multiple organ systems.^47,48^ Although electronic frailty measures such as the HFRS incorporate selected diagnosis codes related to frailty, the HFRS does not directly include several major CHIP-associated outcomes such as coronary artery disease, heart failure, hematologic malignancies, or chronic liver disease. Moreover, the consistent associations across incident frailty, accelerated frailty progression, survey-based frailty, directly measured physical frailty, clinically assessed frailty, and proteomic healthspan profiles argue against diagnosis-code accumulation as the sole explanation and suggest that somatic clonal expansion contributes to a broader decline in physiological resilience during aging.

Second, our findings suggest that the biological consequences of clonal hematopoiesis depend on the characteristics of the clone rather than simply the presence of clonal expansion. This was evident across different classes of somatic alterations, including autosomal mCAs, mLOX, and mLOY. The stronger association of autosomal mCAs with frailty compared with sex chromosome mosaicism may reflect distinct biological consequences. Autosomal mCAs have been linked to broad immune dysfunction, with GWAS loci enriched in immune-cell regulatory regions^49^ and PheWAS analyses implicating a wide range of infectious, inflammatory, and hematologic phenotypes.^50^ In contrast, sex chromosome mosaicism has been associated with distinct molecular and cellular consequences, including altered immune function and tissue remodeling programs.^50,51^ For CHIP, we found that *DNMT3A* R882 hotspot mutations and *TET2* loss-of-function mutations showed stronger associations with frailty than other mutation classes. This pattern is consistent with prior work demonstrating that specific CHIP mutation classes carry distinct clinical and biological consequences^52^. Biologically, this is plausible because driver genes and mutation classes differ in their effects on hematopoietic stem cell fitness, myeloid differentiation, inflammatory signaling, and immune cell output.^53–55^

Third, our findings implicate immune remodeling as one potential pathway linking specific forms of clonal hematopoiesis to frailty. Although frailty has long been associated with immune dysfunction and impaired immune resilience^16,56^, the upstream mechanisms remain incompletely understood. Our findings suggest that immune consequences of clonal hematopoiesis are context- and genotype-dependent rather than uniform across clones. *TET2* loss-of-function mutations showed stronger associations with frailty, and hematopoietic *Tet2* loss-of-function in mice produced frailty-like functional decline accompanied by age-associated T-cell remodeling, in addition to previously described effects on myeloid inflammatory pathways^57,58^. In humans, T-cell activation–related plasma proteins were associated with incident frailty and partially mediated the association between clonal hematopoiesis and frailty. The distinct mediation signatures across CHIP driver genes further suggest subtype-specific immune pathways.

Although CHIP has traditionally been characterized by inflammatory myeloid activation, emerging evidence supports context-dependent effects on immune function, including altered lymphoid responses and immune states^26,54^. Thus, specific driver mutations, cellular compartments, and aging contexts may shape how clonal hematopoiesis influences immune resilience and functional decline.

This study has several limitations. First, the observational human analyses preclude causal inference, and residual confounding by comorbidities, healthcare utilization, and other age-related factors may remain. Second, the primary frailty phenotype was EHR-derived and may be influenced by diagnostic coding and differences in healthcare capture across cohorts. Third, proteomic analyses were restricted to UK Biobank participants with available Olink data, which may limit generalizability. Fourth, clonal hematopoiesis was generally assessed at a single time point, limiting characterization of clonal evolution; in the WHI longitudinal analysis, limited incident events, attrition, and survivor bias may also have reduced statistical power.

In conclusion, across multiple human cohorts and complementary frailty phenotypes, clonal hematopoiesis was consistently associated with greater frailty burden and accelerated functional decline, with substantial heterogeneity across clonal lesions. Experimental and immune profiling findings further support immune dysregulation as a plausible biological link. Together, these results broaden the clinical relevance of clonal hematopoiesis beyond disease-specific risk and position it as a potential contributor to unhealthy aging. Future longitudinal and interventional studies are needed to determine whether CH-informed risk stratification or immune-targeted strategies can preserve physiological reserve and extend healthspan.

## Methods

### Data Availability

Individual-level sequence data, CHIP calls and polygenic scores have been deposited with UK Biobank and are available to approved researchers through application, as done with other genetic datasets to date under data fields (30105, 30106, 30107). The genotypes and phenotypes of UKB and AoU participants are available by application to the UKB (https://www.ukbiobank.ac.uk/register-apply/) and AoU (https://allofus.nih.gov/), respectively. For Vanderbilt BioVU, clonal hematopoiesis of indeterminate potential sequencing calls are available through controlled access to qualified researchers. Due to Vanderbilt BioVU cohort restrictions, access will require a data use agreement with Vanderbilt University Medical Center, which can be facilitated by the corresponding author. WHI data are publicly available through https://www.whi.org/get-started and https://www.whi.org/propose-a-paper.

### Primary biobank cohorts for frailty analyses

This study utilized data from three large-scale cohorts: the Vanderbilt BioVU biobank (United States; enrollment 2007–2022), the NIH All of Us (AoU) Research Program (United States; enrollment 2018–2023), and the UK Biobank (UKB; United Kingdom; enrollment 2006–2010). The three cohorts are described as follows:

1. All of Us: Analyses were conducted using the Controlled Tier Curated Data Repository (CDR) version 8. AoU is a longitudinal U.S. cohort designed to enroll more than one million participants, with an emphasis on populations historically underrepresented in biomedical research. Detailed cohort and genomic data-generation protocols have been described previously.^59,60^ Eligible participants are adults aged ≥18 years residing in the United States or U.S. territories who provide informed consent.
2. UK Biobank: UKB is a population-based cohort of approximately 500,000 participants aged 40–70 years at recruitment between 2006 and 2010.^61^ This study used available whole-genome sequencing (WGS) and whole-exome sequencing (WES) data. WGS was performed using Illumina NovaSeq platforms, with sequencing data subsequently processed using standardized UKB pipelines and reprocessed using DRAGEN v3.7.8.
3. BioVU is a Vanderbilt University Medical Center biobank with linked de-identified electronic health records (EHRs).^62^ Analyses included participants with available WGS data. Sequencing libraries were prepared using the Illumina DNA PCR-Free Prep protocol and sequenced on Illumina NovaSeq 6000 instruments to a mean coverage of approximately 30×. The Vanderbilt University Medical Center Institutional Review Board oversees BioVU and approved this study.

Participants with available genomic data were initially considered (BioVU: 245,540; AoU: 275,679; UKB: 488,175). We excluded individuals with a documented history of hematologic malignancy or other non-neoplastic clonal disorders (ICD codes outlined in **Supplementary Table 1**). Given the low prevalence of clonal hematopoiesis in younger individuals and to ensure comparable age distributions across cohorts, thereby reducing potential age-related and technical confounding, analyses were restricted to participants aged 40–70 years at baseline.

### Clonal hematopoiesis variant calls

For CHIP, putative somatic SNVs and short indels were called with GATK Mutect2 and filtered according to previously established criteria among the three cohorts as we have previously described.^63,64^ Briefly, 74 canonical CHIP genes were screened for potential CHIP mutations using the Mutect2 somatic variant caller (**Supplementary Table 2**).^65^ Variants included in the preliminary dataset met the following criteria: presence in a pre-established list of candidate CHIP variants, total sequencing depth ≥ 20, alternate allele read depth count ≥ 5, and representation in both sequencing directions (i.e., F1R2 ≥ 1 and F2R1 ≥ 1). CHIP mutations were defined as those with a variant allele fraction (VAF) ≥ 0.02. CHIP detection across all cohorts was derived from whole genome/exome sequencing data, using a consistent detection method and the same canonical CHIP driver genes list, which specifies candidate missense and indel variants for each gene, as well as a set of genes in which truncating and splice site variants may be considered.

mCAs were identified using haplotype phasing–based approaches across cohorts. In BioVU, mCA detection from whole-genome sequencing (WGS) data was performed using *MoChA* version 1.11. *MoChA* leverages genotype information to estimate allelic balance and read depth at heterozygous loci, enabling detection of allelic imbalances through deviations in B-allele frequency (BAF) and read coverage. In UKB, mCA detection from genotyping intensity data has been previously described.^66^ In the AoU, mCA detection was performed starting from raw IDAT files generated using a single version of the Infinium Global Diversity Array. *MoChA* version 1.11 was applied using methods harmonized with those implemented in UKB and BioVU. Additional details regarding mCA calling and quality control procedures are provided in our previous publication.^67^ Detected events were classified as copy-neutral loss of heterozygosity (CN-LOH), deletions, duplications, or events with undetermined copy number changes.

### Hospital Frailty Risk Score calculation

In the UK Biobank, All of Us, and BioVU cohorts, frailty was assessed using the Hospital Frailty Risk Score (HFRS), a predefined ICD code-based frailty index derived from routinely collected healthcare data.10 The HFRS was originally developed and validated using hospital administrative data and is based on 109 frailty-associated ICD-10 diagnostic codes, each weighted according to its association with adverse clinical outcomes (**Supplementary Table 3**). ICD codes recorded in the electronic health record were mapped to the corresponding HFRS definitions and harmonized across cohorts prior to analysis. For each participant, HFRS was calculated longitudinally as the cumulative weighted sum of qualifying frailty-related diagnostic codes recorded up to each time point. Baseline HFRS was defined as the cumulative HFRS at the time of blood sampling used for CH ascertainment. During follow-up, HFRS was updated at each subsequent clinical encounter as new qualifying diagnostic codes were recorded. Because the score was cumulative, previously recorded qualifying diagnoses continued to contribute to subsequent HFRS values. For time-to-event analyses, participants with frailty at baseline were excluded from the corresponding analysis. Incident frailty was defined as the first date during follow-up on which the cumulative HFRS crossed the predefined threshold of >5, and severe frailty as the first date on which HFRS crossed >15, consistent with prior validation studies.^10,30^ Thus, the event date corresponded to the date of the clinical encounter at which newly recorded qualifying diagnosis codes first increased the cumulative HFRS above the respective threshold. Participants who did not reach the outcome threshold were censored at the time of last follow-up, death, or the end of the study period, whichever occurred first. To evaluate frailty as a continuous and longitudinal phenotype independent of these categorical thresholds, we additionally modeled trajectories of repeated HFRS measurements over follow-up using linear mixed-effects models.

### Survey-based frailty index

In UK Biobank only, we evaluated whether associations with CHIP extended beyond EHR-derived frailty to complementary measures of deficit accumulation. A survey-based frailty index (FI)^11^ was calculated from 49 self-reported baseline questionnaire items covering general health, diagnosed diseases, disability, and mental health, as previously described. Individual deficits were generally coded as present (1) or absent (0), and the FI was calculated as the number of deficits present divided by the number of non-missing deficits assessed. Higher FI values indicated greater frailty. Participants missing more than 10 of the 49 items were excluded.

### Healthspan Proteomic Score

The Healthspan Proteomic Score (HPS)^33^ was calculated among participants with available baseline plasma proteomic data using the previously published algorithm. The score incorporates chronological age and plasma levels of 86 proteins to estimate the 10-year probability of remaining free from healthspan-ending events. Missing protein measurements were imputed using k-nearest-neighbor imputation with k = 10, implemented in the multiUS package in R. GLIPR1, NPM1, and PCOLCE were excluded from imputation because of their high missingness rates. The published Gompertz model was then applied to estimate each participant’s 10-year risk of reaching the end of healthspan, defined by the first occurrence of cancer, diabetes, heart failure, myocardial infarction, stroke, chronic obstructive pulmonary disease, dementia, or death. HPS was calculated as 1 minus the predicted 10-year risk, such that higher values indicated a greater predicted probability of maintaining healthspan. For descriptive analyses, low HPS was defined as a score ≤0.73.

### Measured frailty assessment in Women’s Health Initiative

As a complementary analysis, we examined participants from the Women’s Health Initiative (WHI) with directly measured frailty phenotypes and CHIP assessment at the WHI baseline examination and the Long Life Study (LLS) visit. A total of 3,997 participants had complete data for all frailty components and were included in the measured frailty analyses. Frailty assessment in WHI has been previously described.^68^ Frailty was defined using five components: slow gait speed, weak grip strength, low physical activity, exhaustion, and weight loss. Slow gait speed and weak grip strength were defined as the lowest 20th percentile after BMI adjustment. Physical activity was quantified as total recreational energy expenditure (MET-hours/week), with the lowest quintile classified as low activity. Exhaustion was assessed using the RAND-36 energy/fatigue scale, and participants in the lowest quintile were classified as exhausted. Weight loss was defined based on self-reported recent weight loss on Form 155. Each component was coded as present (1) or absent (0) and summed to produce a frailty score ranging from 0 to 5. Consistent with the Fried phenotype, frailty was defined as a score ≥3.^12^

Peripheral blood DNA collected at both time points was analyzed using high-depth targeted sequencing (>4,000× depth) of established clonal hematopoiesis driver genes, as previously described^69^. CHIP was defined as the presence of a qualifying somatic driver mutation with a variant allele fraction (VAF) ≥2%, and large CHIP was defined as VAF >10%. The availability of targeted deep-sequencing data at both baseline and LLS enabled evaluation of CHIP according to the time point relevant to each analysis. For longitudinal analyses, CHIP status at the WHI baseline examination was evaluated in relation to incident frailty at LLS among participants who were nonfrail at baseline. For cross-sectional analyses, CHIP status and frailty were assessed contemporaneously at the LLS visit.

### Clinical frailty scale in three human aging cohorts

Participant samples and data from three previously completed human aging cohort studies conducted between 2009 and 2018 were included in the analysis^35^ (n=205; **Supplementary Table 9**). These included adults aged 65 and older residing in the communities surrounding Greater Sudbury (Ontario, Canada), Vancouver (British Columbia, Canada) and Hartford (Connecticut, USA). For only one study were some participants recruited because of a chronic condition (i.e. diabetes), while the remaining represent a convenience sample from the community. Peripheral blood mononuclear cells were obtained pre- and post-vaccination and stored in vapour phase liquid nitrogen until DNA extraction for the current study. CHIP was assessed using targeted next-generation sequencing of peripheral blood DNA with custom CHIP gene panels, achieving high sequencing depth (>1,500×).^70^ The CHIP screening workflow and variant filtering criteria were consistent with our primary analysis. Age and sex were available for all participant samples, along with a pre-vaccination assessment of the Clinical Frailty Scale (CFS)^71^. A cumulative link mixed model (CLMM) was fitted using the *clmm* function from the R *ordinal* package to evaluate the association between CHIP status and frailty severity. The CFS was modeled as an ordered categorical outcome. Models included CHIP status as the primary exposure, age and sex as fixed effects, and random intercepts for cohort and calendar year to account for clustering and temporal variation.

### Statistical Analysis

Descriptive statistics were used to summarize baseline characteristics. Continuous variables are reported as medians and interquartile ranges, and categorical variables as counts and percentages. The date of blood sampling used for genomic assessment was defined as time zero for all longitudinal analyses. Associations of clonal hematopoiesis with incident frailty were assessed separately within each cohort using Cox proportional hazards models. Participants with prevalent frailty at baseline (HFRS >5) were excluded from analyses of incident frailty. Participants were followed from the date of blood sampling until the first occurrence of the corresponding frailty outcome, death, the last available clinical record, or the end of the study period, whichever occurred first. CHIP was analyzed overall and according to the major driver genes *DNMT3A*, *TET2*, *ASXL1*, and *JAK2*. mCAs were evaluated as autosomal mCAs, mLOX, and mLOY. CHIP was additionally evaluated using a variant allele fraction (VAF) threshold of ≥10% to assess associations with larger clones relative to non-carriers.

Longitudinal frailty progression was evaluated using repeated frailty index measurements obtained across study visits. Linear mixed-effects models with participant-specific random intercepts were used to account for within-person correlation among repeated measurements. Age at each visit was used as the longitudinal time scale. Models included clonal hematopoiesis status, standardized age at visit, and their interaction term. The interaction coefficient represented the difference in the rate of age-related frailty progression between carriers and non-carriers per 1-standard-deviation increase in age. Models were additionally adjusted for sex, smoking status, and genetic ancestry principal components 1***-***5.

Cox models were adjusted for age at blood sampling, age squared, sex, smoking status, and genetic ancestry principal components 1-5. In sensitivity analyses, Cox models were further adjusted for baseline HFRS as a continuous variable to account for residual differences in frailty burden among participants without prevalent frailty. Cohort-specific estimates were combined using inverse-variance–weighted meta-analysis. Hazard ratios from Cox models were pooled on the log scale and subsequently exponentiated, whereas regression coefficients from linear mixed-effects models were pooled directly. Between-cohort heterogeneity was assessed using Cochran’s Q test and the I^2^ statistic. Fixed-effects meta-analysis was used when I^2^ was <50%, whereas random-effects meta-analysis was used when I^2^ was ≥50%. All statistical tests were two-sided, and *P* <0.05 was considered statistically significant unless otherwise specified.

### Mouse model

All the animal care and used protocols were approved by the McMaster Animal Research Ethics Board (no. 17-05-19) and Home Office (University of Aberdeen, PP2736251) and performed in accordance with Animal Care guidelines. *Tet2^f/f^* (B6;129S-Tet2tm1.1Iaai/J) and Vav1-iCre (B6.Cg-Tg(Vav1-icre)A2Kio/J) mice were bred at Queen’s University and the University of Aberdeen to produce *Tet2^f/f^ Vav1-icre^-/-^*(*Tet2^f/f^*) and *Tet2^f/f^ Vav1-icre^+/-^*(*Tet2^-/-^*) genotypes (exon 3 of Tet2 gene is floxed) in the hematopoietic system. For all analyses, both male and female mice were used. Mice were maintained under a 12-h light-dark cycle at 22 ±2°C and 55 ± 5% air humidity. Mice had ad libitum access to Teklad irradiated global 14% protein diet (Cat. no. 2914, Envigo) and autoclaved reverse osmosis water.

4 to 6-month-old *Tet2^-/-^* and *Tet2^f/f^*were placed in a multiplexed metabolic phenotyping system (Promethion, Sable Systems International) and allowed to acclimatize for 24 hours. Following acclimation, continuous assessment of aerobic metabolism (VO_2_, VCO_2_, and RER), food and water intake, body weight, and physical locomotor activity were taken for an additional 48 to 60 hours. We adapted the mouse clinical frailty index developed by Whitehead et al.^36^, which scores clinical indicators of aging grouped into six functional categories: integument, physical/musculoskeletal, ocular/nasal, digestive/urogenital, respiratory, and discomfort. Each parameter is rated on a three-point scale (0 = healthy; 0.5 = mild-to-moderate impairment, with the mouse showing some impact; 1 = severe impairment with clear impact on well-being), and an overall frailty score is calculated for each animal by summing scores across all parameters.

Using this index, we assessed phenotypic signs of aging in middle aged (12-14 month old) mice in the following categories: integument (alopecia, dermatitis, whisker loss, coat condition); physical/musculoskeletal (tumors, distended abdomen, kyphosis, tail stiffening, gait disorders, tremor, forelimb grip strength, body condition); ocular/nasal (cataracts, corneal opacity, ocular discharge, microphthalmia, vision loss, menace reflex, nasal discharge); digestive/urogenital (rectal prolapse, vaginal/uterine/penile prolapse, diarrhea); respiratory (rate and depth of breathing); and discomfort (mouse grimace scale score, piloerection), together with body weight. Forelimb muscle strength was measured using a grip-strength meter, and motor coordination and fatigue resistance were assessed using a rotarod test and a cage-hang test.

### Proteomic Analyses in humans

Proteomic profiling was performed in 52,705 UK Biobank participants using the Olink proximity extension assay platform (Olink Proteomics, Uppsala, Sweden), which quantified 1,465 circulating proteins across multiple biological panels, including cardiovascular, inflammation, cardiometabolic, neurology, and oncology panels. To investigate immune mechanisms linking clonal hematopoiesis to frailty, we focused specifically on proteins annotated under the Gene Ontology term GO:0042110 (T cell activation), given established evidence implicating adaptive immune remodeling and chronic inflammatory signaling in clonal hematopoiesis and age-related functional decline. A total of 148 T cell activation–related proteins were evaluated.

Associations between circulating protein levels and incident frailty were assessed using Cox proportional hazards models, with protein levels modeled as continuous exposures. Models were adjusted for age at blood draw, age squared, sex, smoking status, and genetic ancestry principal components. Multiple testing correction was applied using a Bonferroni-adjusted threshold based on the number of proteins tested (*P* < 0.05/148). To assess whether T cell activation–related proteins mediated the association between clonal hematopoiesis and frailty, we conducted mediation analyses using the R package *mediation*. This framework decomposes the total effect of CHIP on frailty into a direct effect (independent of protein levels) and an indirect effect mediated through circulating protein levels. A two-stage regression approach was implemented. First, a mediator model was fitted to estimate the association between CHIP status and protein levels, adjusting for age at blood draw, age squared, sex, smoking status, and genetic ancestry principal components. Second, an outcome model was fitted to estimate the association between protein levels and incident frailty while adjusting for CHIP and the same covariates. The average causal mediation effect (ACME), average direct effect (ADE), and proportion mediated were estimated. Statistical inference was obtained using nonparametric bootstrapping. FDR correction was applied to account for multiple testing across proteins.

### Code Availability

The code is publicly available and can be found at https://github.com/bicklab/frailty.

## Data Availability

All data produced in the present study are available upon reasonable request to the authors

https://github.com/bicklab/frailty

## Acknowledgments

A.G.B. is supported by National Institutes of Health (DP5OD029586, R01AG088657, and R01AG083736); a Career Award for Medical Scientists from the Burroughs Wellcome Fund; a Pew-Stewart Scholar for Cancer Research award (supported by the Pew Charitable Trusts and the Alexander and Margaret Stewart Trust); a Hevolution/AFAR New Investigator Award in Aging Biology and Geroscience Research. L.M.E is supported by Hevolution/AFAR HEV∼NI23013 and NIA R01AG046938. G.I.B was supported by the Interdisciplinary Quantitative Biology Program and Hevolution/AFAR HEV∼NI23013. C.Q is supported by awards from Friends of ANCHOR and AMS Springboard, and the generous philanthropic support of Dr Hugh Young Rienhoff, Jr. E.A.W was supported by a grant from the National Heart, Lung, and Blood Institute, National Institutes of Health (R01HL148565). M.J.R is supported by a Canadian Institutes of Health Research (CIHR) Project Grant (PJT-175032). Y.P was supported by a grant from the National Institutes of Aging (F30 AG099331). The WHI program is funded by the National Heart, Lung, and Blood Institute, National Institutes of Health, U.S. Department of Health and Human Services through contracts 75N92021D00001, 75N92021D00002, 75N92021D00003, 75N92021D00004, 75N92021D00005. Human aging cohorts were derived from participants enrolled in an influenza vaccine trial (R01 AG048023). A list of WHI Investigators is available at https://www.whi.org/doc/WHI-Investigator-Long-List.pdf.

We gratefully acknowledge the late Dr. Janet McElhaney, principal investigator of the human aging cohorts, for her leadership in establishing these cohorts and for the enduring scientific resource they provide.

## Extended Data Figure

**Extended Data Fig 1.**
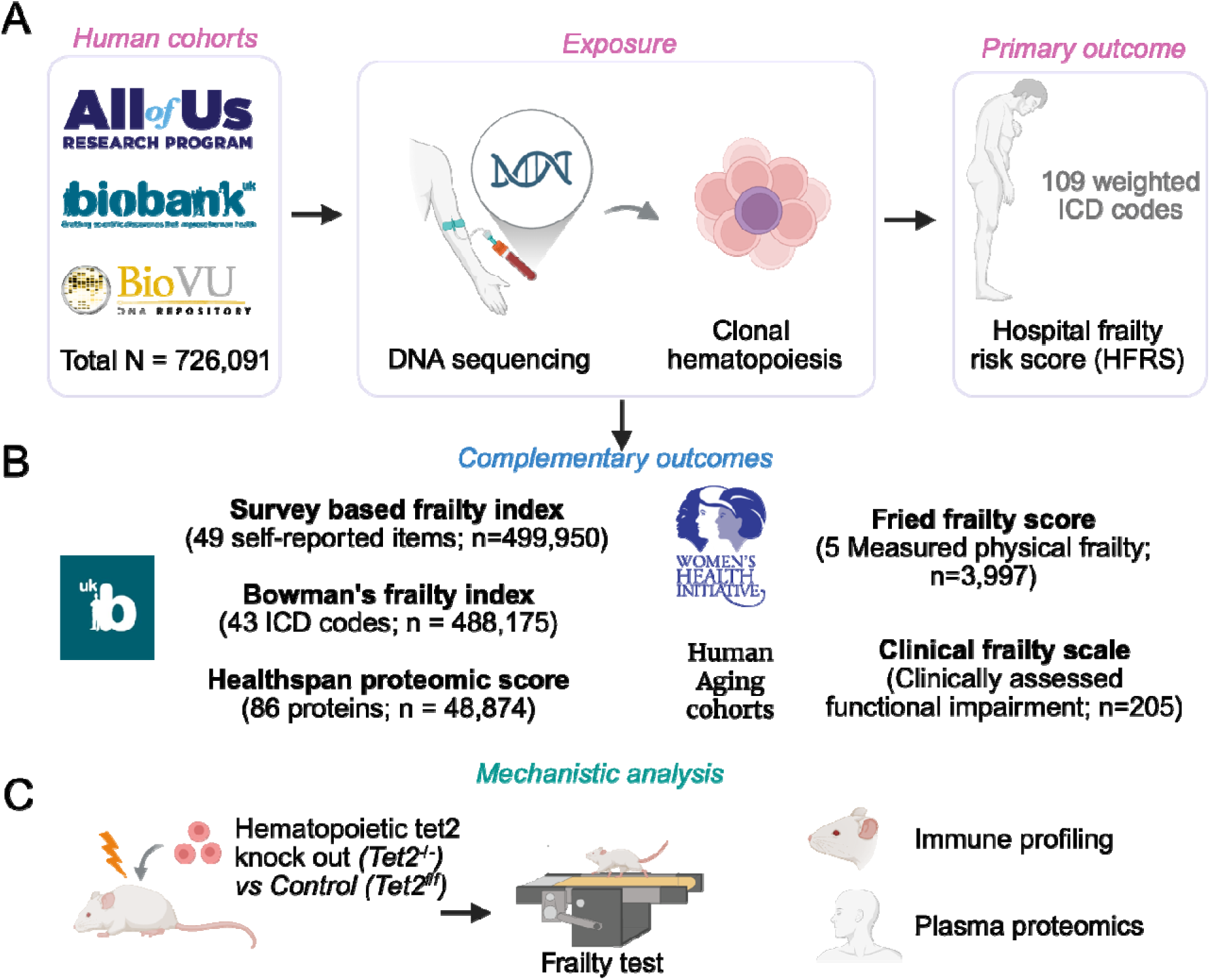
Study design and analytic framework. (A) Primary study design across three large human biobanks, including the All of Us Research Program, UK Biobank, and Vanderbilt BioVU. Clonal hematopoiesis was identified from DNA sequencing data and evaluated in relation to the primary outcome of incident frailty, defined using the Hospital Frailty Risk Score (HFRS), an electronic health record–derived score based on 109 weighted ICD codes. (B) Complementary frailty and healthy-aging outcomes were evaluated to assess the robustness and generalizability of the primary findings. These included a survey-based 49-item frailty index, Bowman’s electronic frailty index, and the Healthspan Proteomic Score in UK Biobank; a standardized measure of physical frailty in the Women’s Health Initiative and clinically assessed functional impairment in human aging cohorts. (C) Mechanistic and functional analyses included a hematopoietic *Tet2* loss-of-function mouse model to assess frailty-like functional decline and immune remodeling, together with human immune profiling analyses based on plasma proteomics.

**Extended Data Fig 2.**
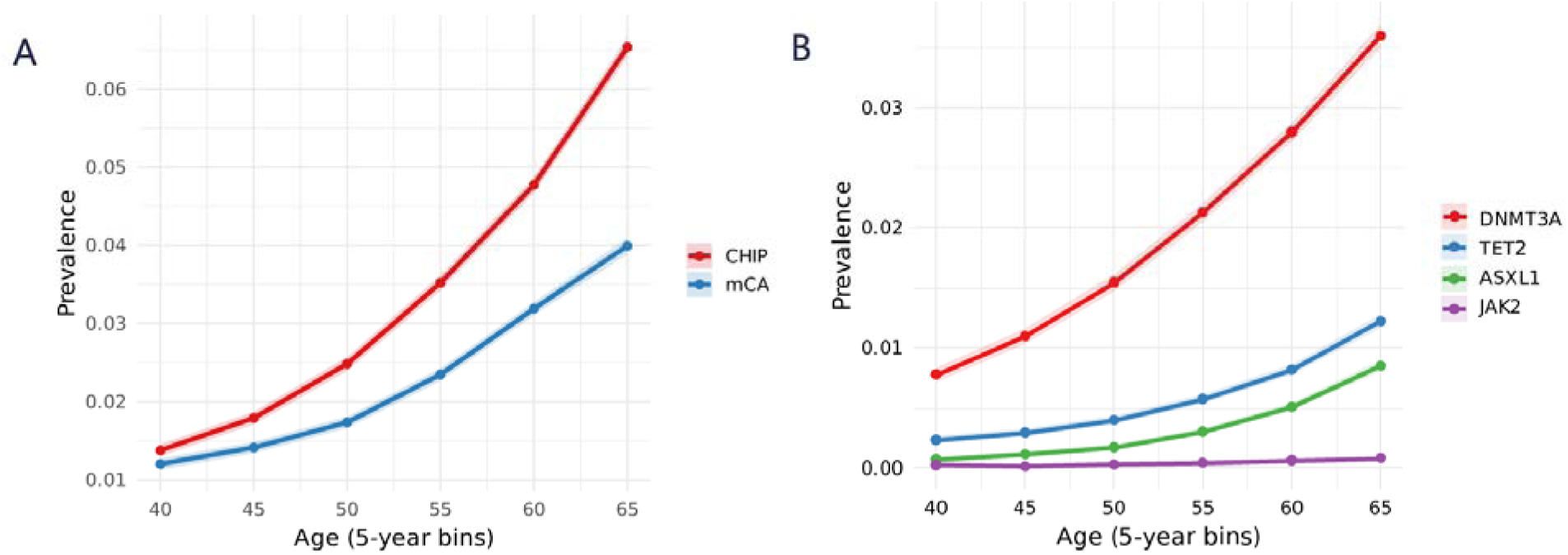
Age-associated increase in clonal hematopoiesis and mosaic chromosomal alterations. (A) Prevalence of overall CHIP and mCA across 5-year age bins. (B) Age-specific prevalence of individual CHIP driver genes, including *DNMT3A, TET2, ASXL1,* and *JAK2*. Lines represent observed prevalence within each age group, and shaded areas indicate 95% confidence intervals. CHIP and mCA prevalence increased progressively with age, with *DNMT3A* being the most common driver mutation across all age groups.

**Extended Data Fig 3.**
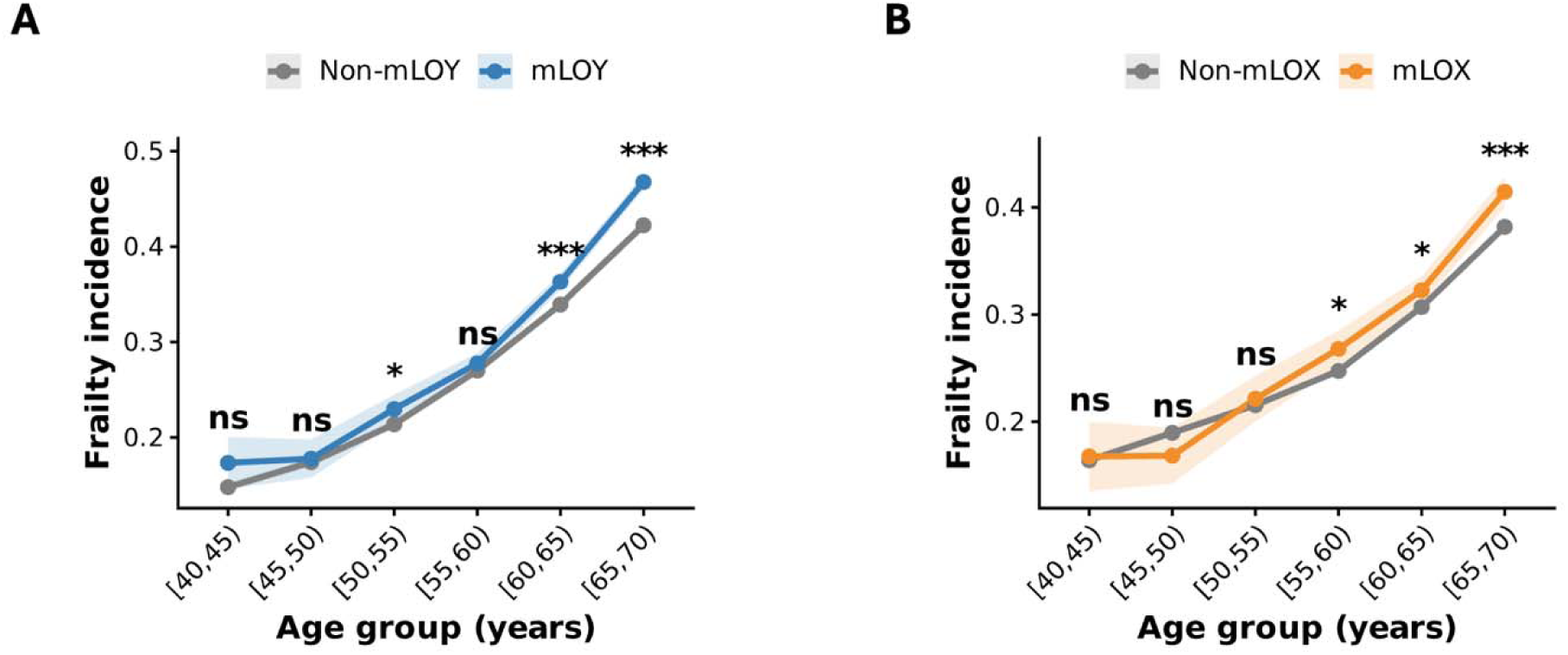
Age-specific frailty incidence according to mosaic loss of chromosome Y or X. **(A)** Age-specific frailty incidence among males with and without mosaic loss of chromosome Y (mLOY). **(B)** Age-specific frailty incidence among females with and without mosaic loss of chromosome X (mLOX). Points and connecting lines indicate frailty incidence within 5-year age groups, and shaded bands indicate 95% confidence intervals.

**Extended Data Fig 4.**
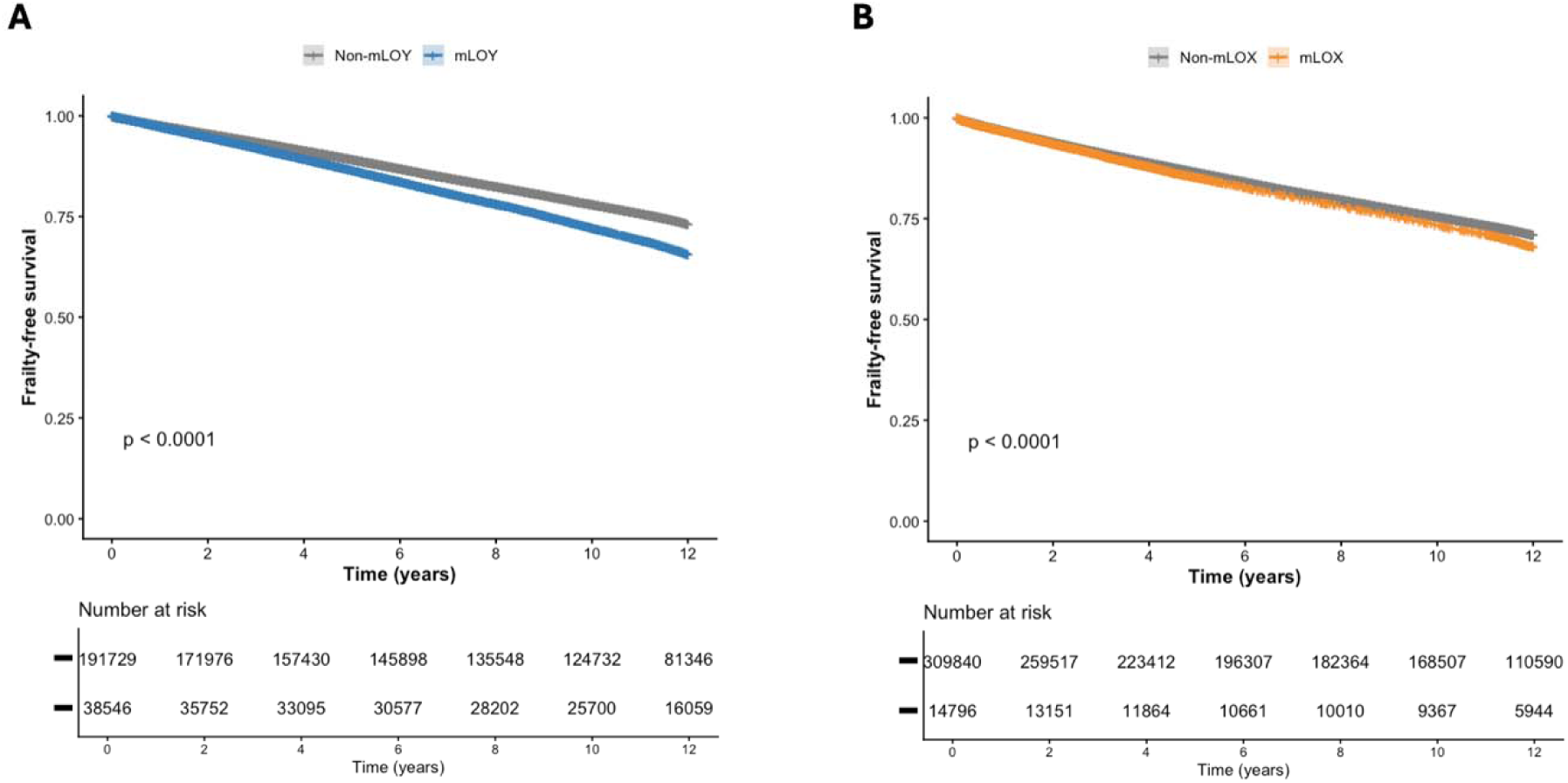
Frailty-free survival according to mosaic loss of chromosome Y or X. Kaplan–Meier curves show frailty-free survival among **(A)** males with and without mosaic loss of chromosome Y (mLOY) and **(B)** females with and without mosaic loss of chromosome X (mLOX). Numbers at risk are shown below each panel. Unadjusted frailty-free survival differed between carriers and non-carriers for both mLOY and mLOX (log-rank *P* < 0.0001), although the absolute separation between curves was modest.

**Extended Data Fig 5.**
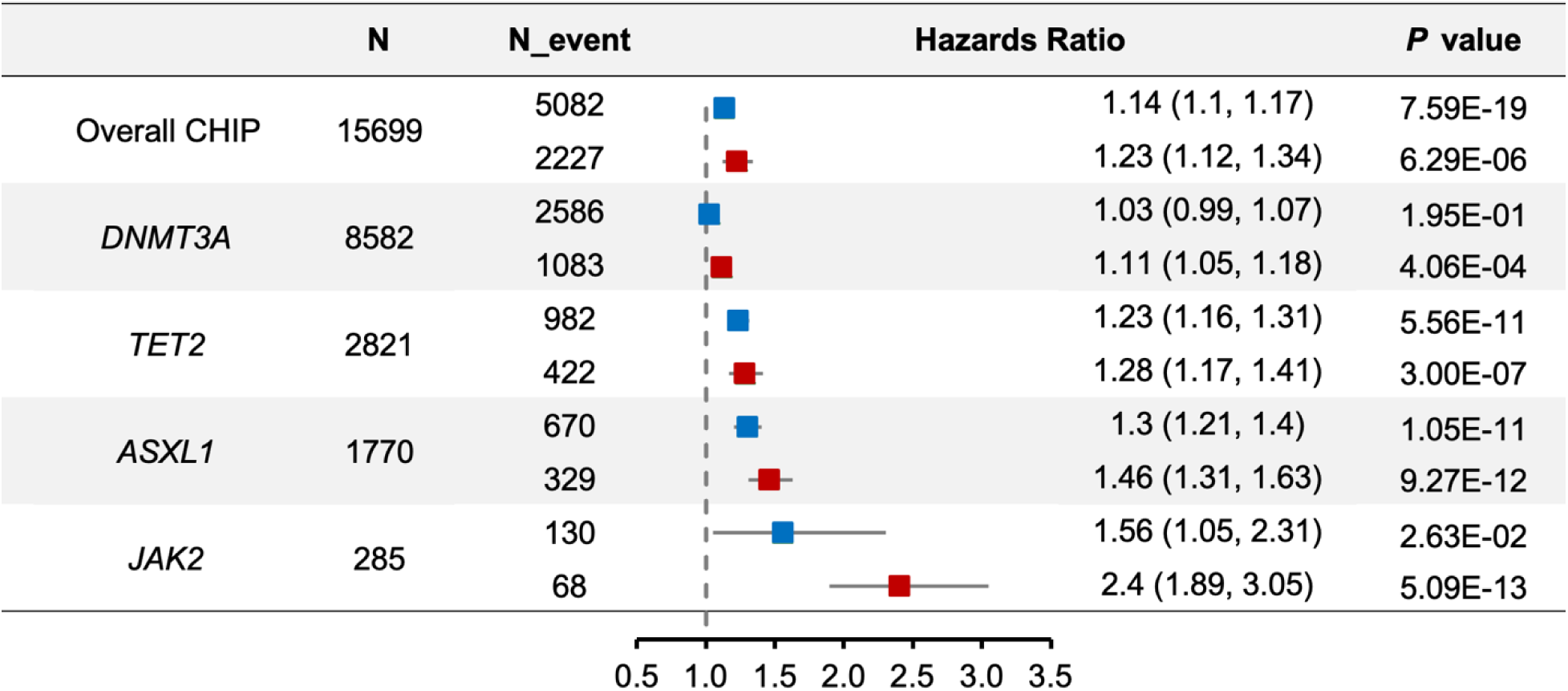
Associations of large CHIP (VAF >10%) with incident frailty and severe frailty. Forest plot showing hazard ratios (HRs) and 95% confidence intervals for incident frailty (blue squares) and severe frailty (red squares) among individuals with large clonal hematopoiesis (variant allele fraction >10%). Estimates were derived from Cox proportional hazards models adjusted for age, age^2^, sex, smoking status, and genetic principal components (PC1–PC5), and combined using inverse-variance–weighted meta-analysis across cohorts, with fixed- or random-effects models selected according to between-cohort heterogeneity. Numbers of carriers (N), Numbers of events among carriers (N_event), hazard ratios with 95% confidence intervals, and corresponding P values are shown.

**Extended Data Fig 6.**
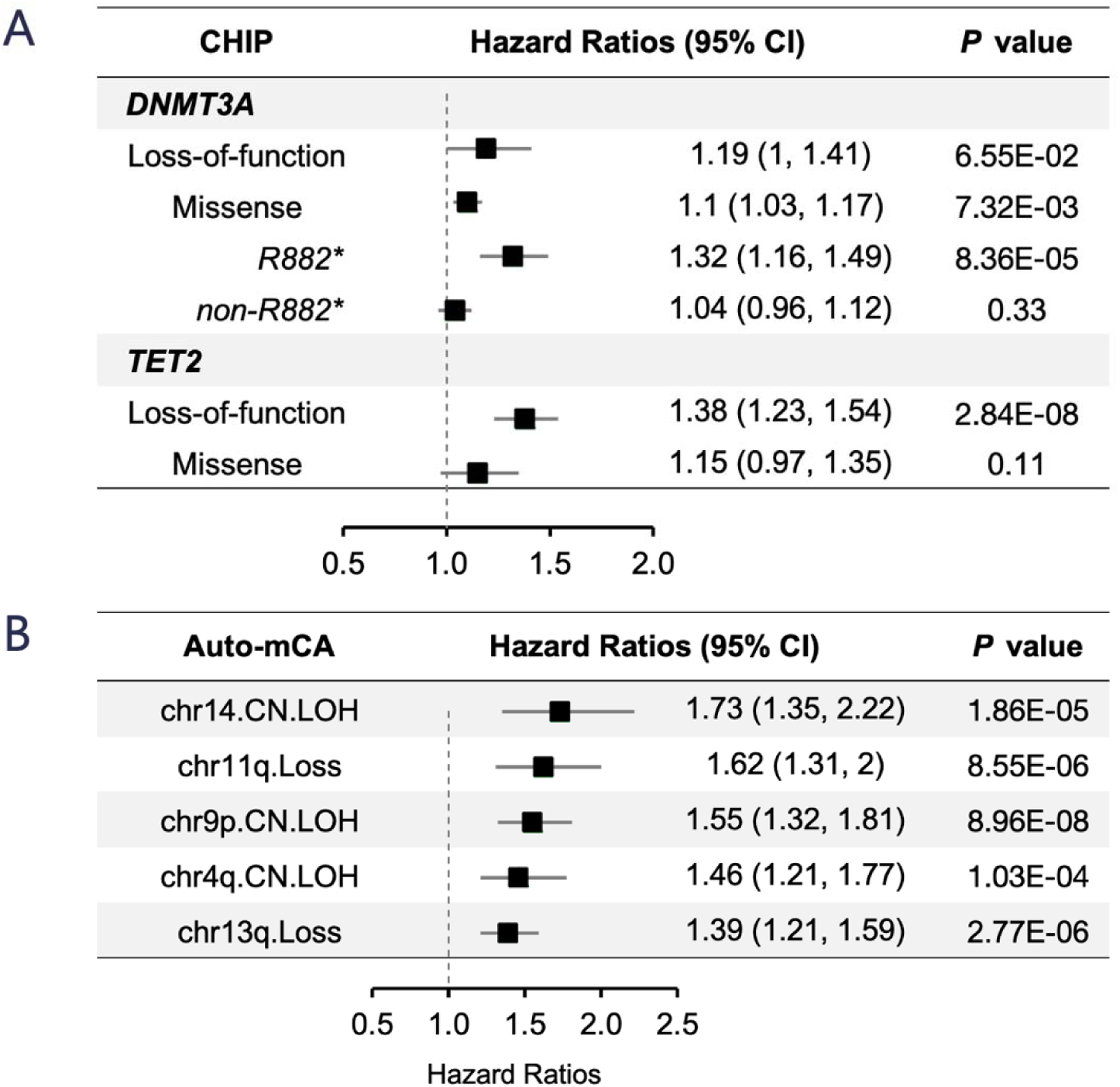
Subtype-specific associations of clonal hematopoiesis mutations with incident frailty. Forest plots display hazard ratios (HRs) and 95% confidence intervals derived meta-analyses across UK Biobank, All of Us, and BioVU cohorts. Cohort-specific Cox proportional hazards models were adjusted for age, age2, sex, smoking status, and genetic principal components (PC1-PC5). **(A)** Mutation subtype analysis of *DNMT3A* and *TET2*-CHIP. **(B)** Chromosome arm-specific analysis of autosomal mCAs. Five recurrent events were significantly associated with increased frailty incidence, including chr14 copy-neutral loss of heterozygosity (CN-LOH), chr11q loss, chr9p CN-LOH, chr4q CN-LOH, and chr13q loss. Hazard ratios with corresponding P values are shown. Statistical significance for autosomal mCA subtypes was defined using Bonferroni correction for 181 tests (*P* < 2.76 × 10^-4^).

**Extended Data Fig 7.**
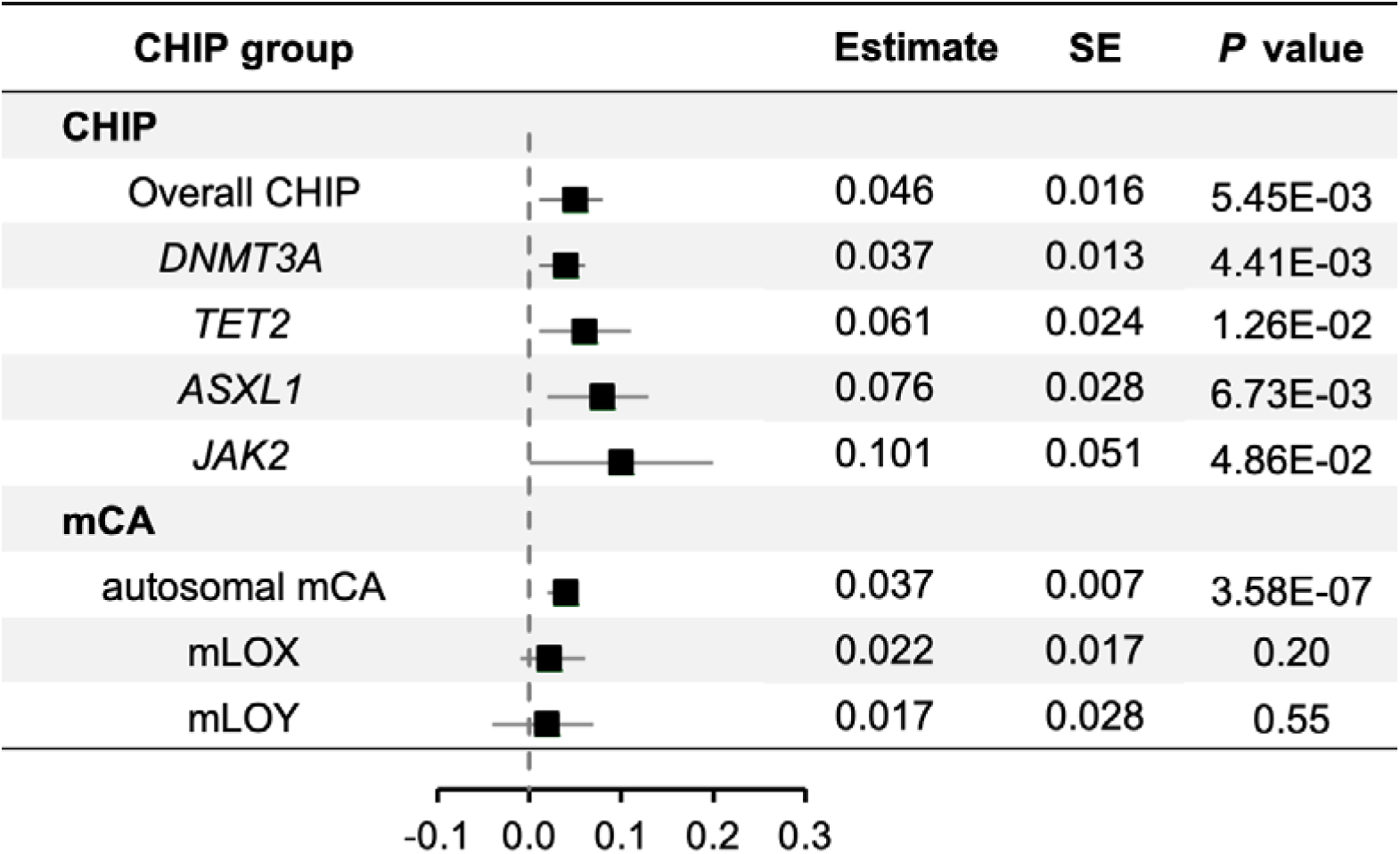
Associations of clonal hematopoiesis with longitudinal frailty trajectory. Estimates were derived from cohort-specific longitudinal linear mixed-effects models and subsequently combined using meta-analysis across UK Biobank, All of Us, and BioVU. Within each cohort, frailty trajectory was modeled using linear mixed-effects regression with random intercepts for participants to account for repeated measures over time. Models included CHIP or mCA status, time, and their interaction term, adjusted for age, age2, sex, smoking status, and genetic principal components (PC1–PC5). The reported estimates represent the interaction term between CHIP/mCA status and time, reflecting differences in frailty progression rate.

**Extended Data Fig 8.**
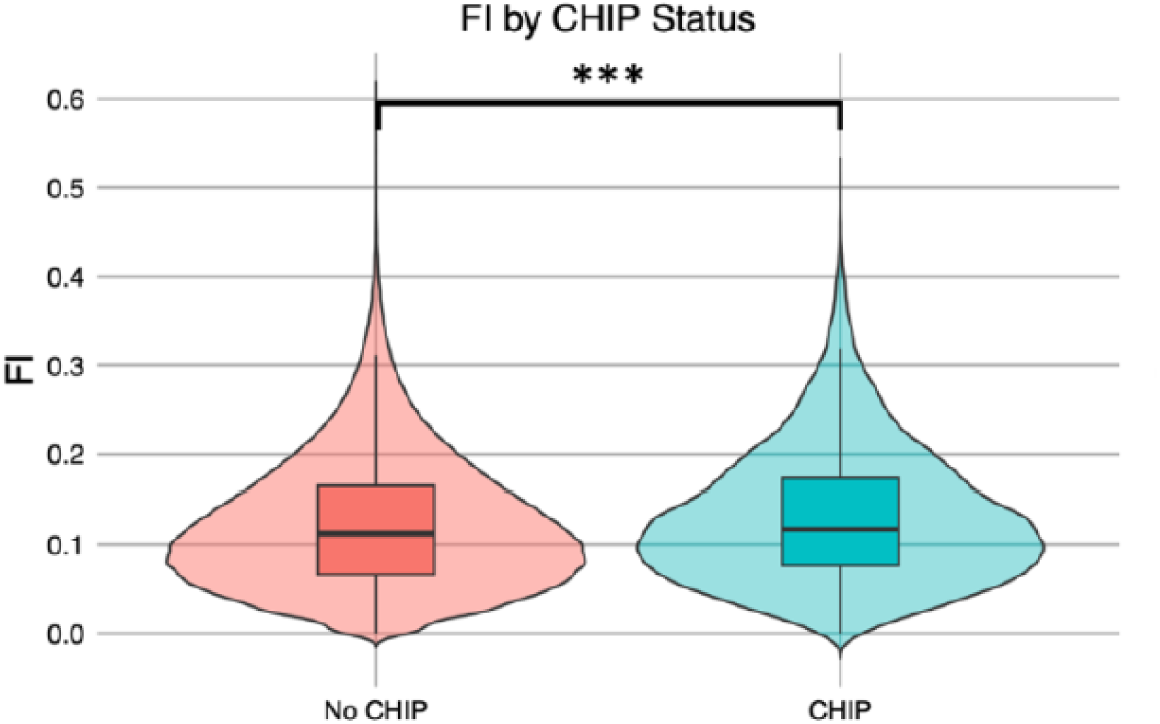
CHIP is associated with higher survey-based frailty index values in UK Biobank. Violin plots show the distribution of baseline frailty index values derived from 49 self-reported health deficits among participants with and without CHIP. Embedded box plots indicate the median and interquartile range. CHIP carriers had higher frailty index values than non-carriers (\*\*\**P* < 0.001).

**Extended Data Fig 9.**
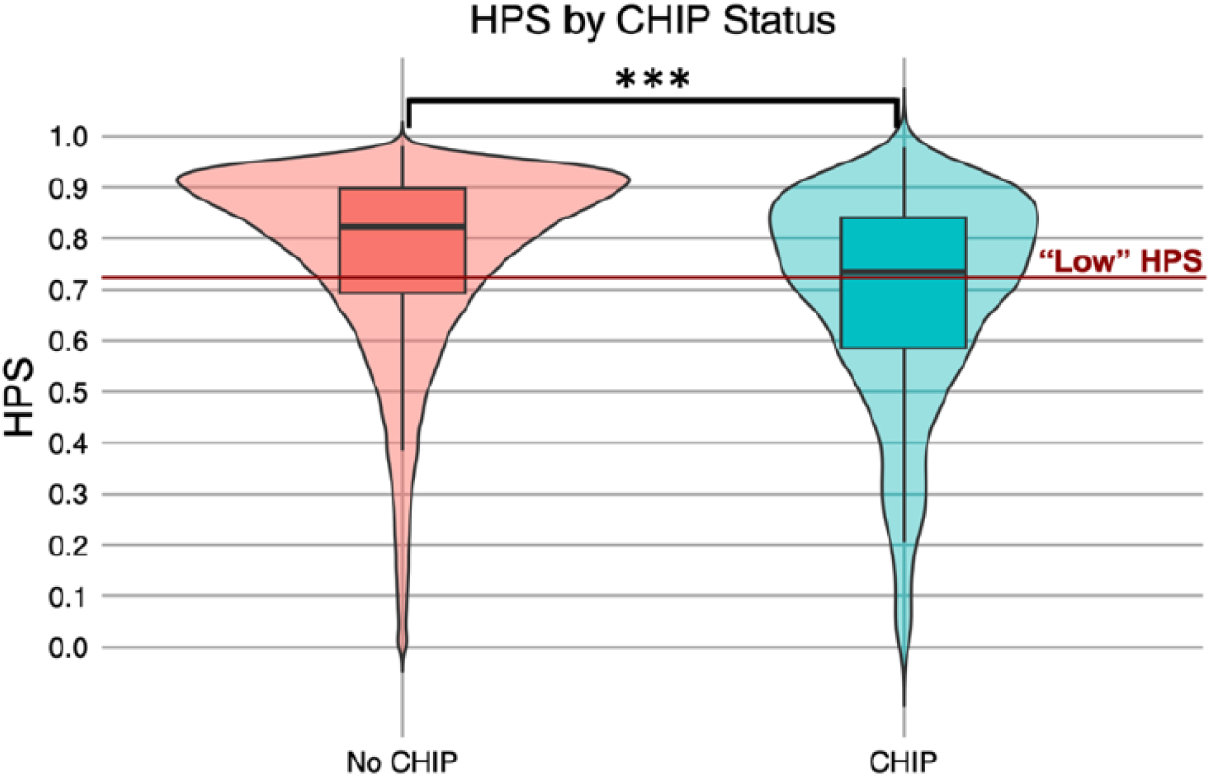
CHIP is associated with lower proteomic healthspan scores in UK Biobank. Violin plots show the distribution of baseline Healthspan Proteomic Score (HPS) values among 48,874 participants with and without CHIP. Embedded box plots indicate the median and interquartile range. CHIP carriers had lower HPS values than non-carriers (median, 0.74 versus 0.82; *P* < 0.001), indicating a lower predicted probability of remaining free from healthspan-ending conditions. The red horizontal line indicates the prespecified threshold for low HPS (≤0.73).

**Extended Data Fig 10.**
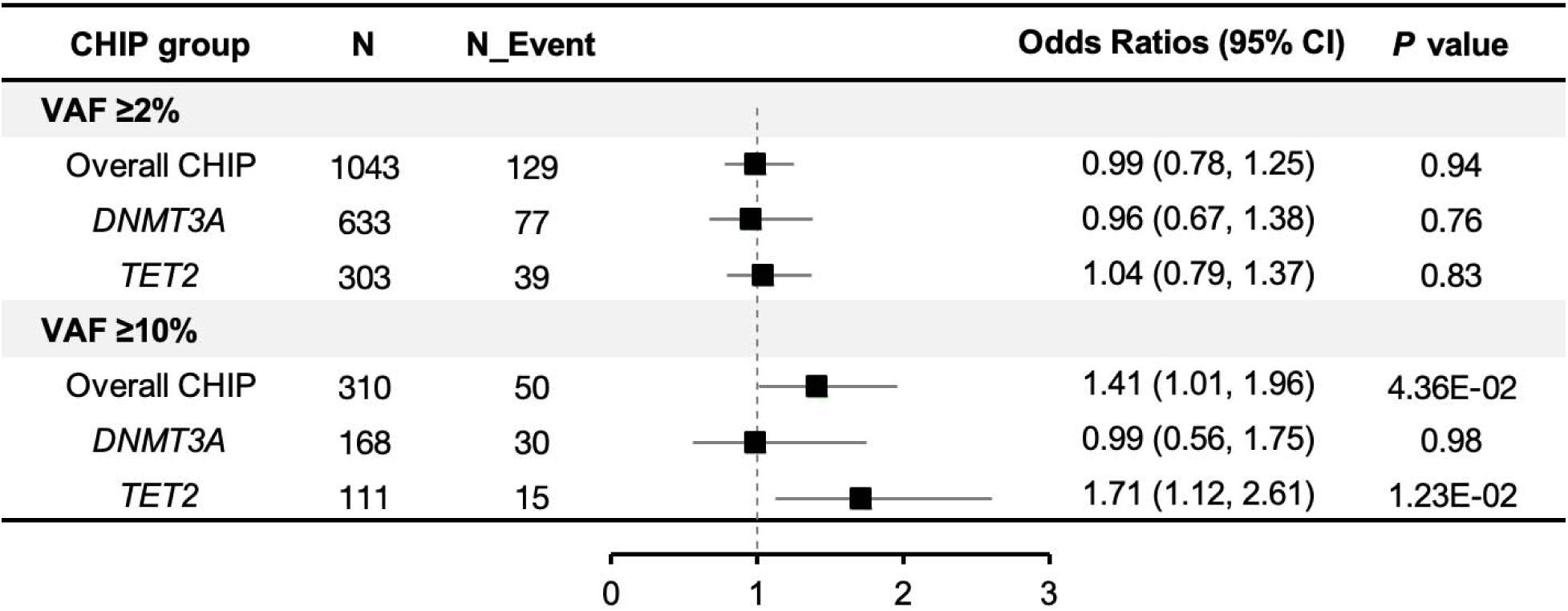
Association of CHIP With Directly Measured Frailty in the Women’s Health Initiative Long Life Study. Forest plot showing odds ratios and 95% confidence intervals for the association between CHIP and prevalent frailty in the Women’s Health Initiative Long Life Study. Frailty was assessed using the Fried phenotype rather than ICD-based definitions. Analyses were stratified by clonal burden using variant allele fraction thresholds of ≥2% and ≥10%. Overall CHIP and driver gene– specific CHIP groups are shown. Odds ratios were estimated using logistic regression models adjusted for age, race, smoking status, and body mass index.

**Extended Data Fig 11.**
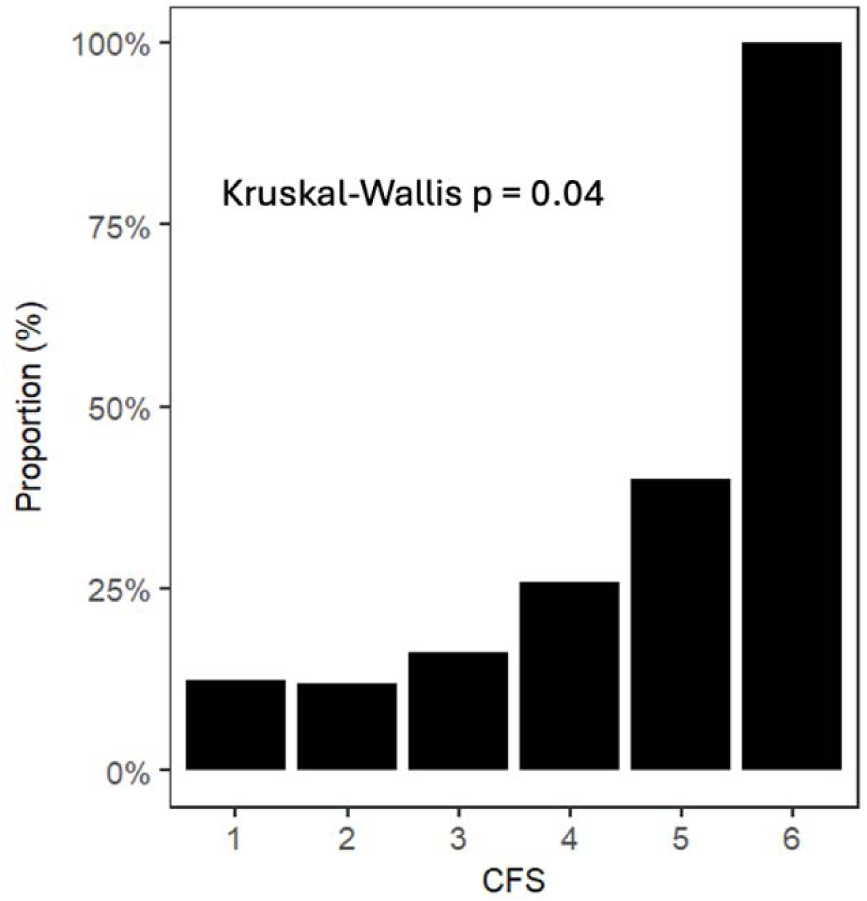
Distribution of CHIP prevalence across Clinical Frailty Scale categories. The proportion of individuals with CHIP is shown across Clinical Frailty Scale (CFS) categories (1–6) among older adults with available CFS assessments. Differences in CHIP prevalence across CFS categories were evaluated using the Kruskal–Wallis test (P = 0.04).

**Extended Data Fig 12.**
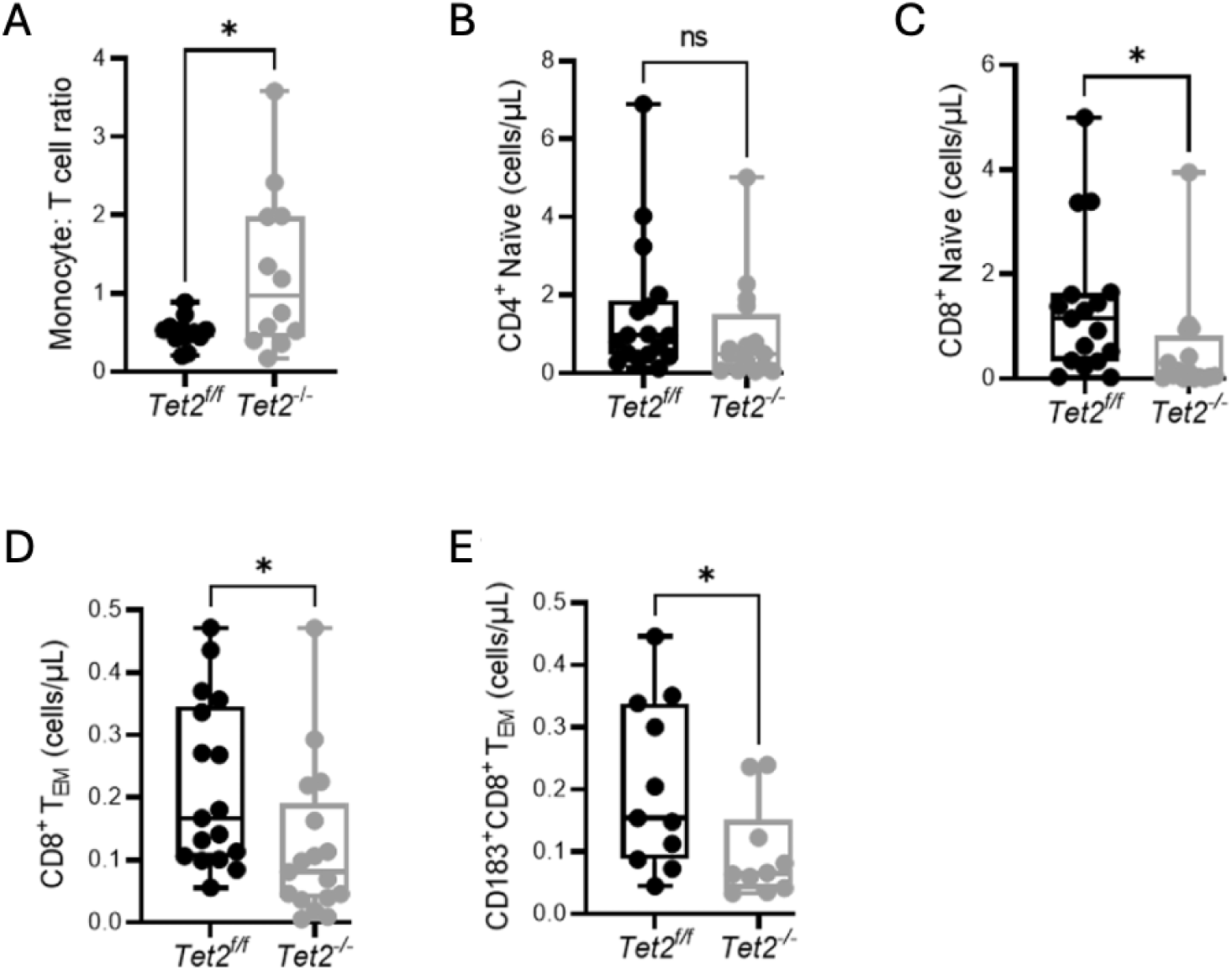
Hematopoietic *Tet2* loss-of-function alters circulating immune-cell composition and T-cell subsets. **(A)** Monocyte-to-T-cell ratio was increased in *Tet2^-/-^* mice compared with *Tet2^f/f^* controls. **(B)** Absolute numbers of naïve CD4^+^ T cells were not significantly different between genotypes. **(C)** Absolute numbers of naïve CD8^+^ T cells were reduced in *Tet2^-/-^* mice. **(D,E)** Circulating CD8^+^ effector-memory T-cell and CD183^+^CD8^+^ effector-memory T-cell numbers were also reduced in *Tet2^-/-^* mice. Data are shown as box-and-whisker plots extending from minimum to maximum, with center lines indicating medians; each point represents one mouse. Comparisons were performed using two-sided Mann–Whitney tests. *P* ≤ 0.05; ns, not significant.

## References

1. Prince, M. J. et al. The burden of disease in older people and implications for health policy and practice. Lancet 385, 549–562 (2015).

2. Chatterji, S., Byles, J., Cutler, D., Seeman, T. & Verdes, E. Health, functioning and disability in older adults – current status and future implications. (2016).

3. Rudnicka, E. et al. The World Health Organization (WHO) approach to healthy ageing. Maturitas 139, 6–11 (2020).

4. Garmany, A. & Terzic, A. Global Healthspan-Lifespan Gaps Among 183 World Health Organization Member States. JAMA Netw Open 7, e2450241 (2024).

5. Beard, J. R. et al. The World report on ageing and health: a policy framework for healthy ageing. The Lancet 387, 2145–2154 (2016).

6. Jang, J., Jung, H., Shin, J. & Kim, D. H. Assessment of Frailty Index at 66 Years of Age and Association With Age-Related Diseases, Disability, and Death Over 10 Years in Korea. JAMA Netw Open 6, e2248995 (2023).

7. Kim, D. H. & Rockwood, K. Frailty in Older Adults. N Engl J Med 391, 538–548 (2024).

8. Clegg, A., Young, J., Iliffe, S., Rikkert, M. O. & Rockwood, K. Frailty in elderly people. The Lancet 381, 752–762 (2013).

9. Yu, Y., Guo, H., Li, Z., Lv, Z. & Chen, Y. Frailty-related multiple health outcomes in older individuals: an umbrella review of systematic reviews and meta-analyses. Age Ageing 55, afag145 (2026).

10. Gilbert, T. et al. Development and validation of a Hospital Frailty Risk Score focusing on older people in acute care settings using electronic hospital records: an observational study. The Lancet 391, 1775–1782 (2018).

11. Williams, D. M., Jylhävä, J., Pedersen, N. L. & Hägg, S. A Frailty Index for UK Biobank Participants. The Journals of Gerontology: Series A 74, 582–587 (2019).

12. Fried, L. P. et al. Frailty in Older Adults: Evidence for a Phenotype. The Journals of Gerontology Series A: Biological Sciences and Medical Sciences 56, M146–M157 (2001).

13. Rockwood, K. A global clinical measure of fitness and frailty in elderly people. Canadian Medical Association Journal 173, 489–495 (2005).

14. Kravvariti, E. et al. Geriatric Frailty Is Associated With Oxidative Stress, Accumulation, and Defective Repair of DNA Double-Strand Breaks Independently of Age and Comorbidities. The Journals of Gerontology: Series A 78, 603–610 (2023).

15. Andersson, R., Mejia-Ramirez, E. & Florian, M. C. Haematopoietic ageing in health and lifespan. Nat Cell Biol 27, 1398–1410 (2025).

16. Tran Van Hoi, E., et al. Biomarkers of the ageing immune system and their association with frailty – A systematic review. Experimental Gerontology 176, 112163 (2023).

17. Ahmad, H., Jahn, N. & Jaiswal, S. Clonal Hematopoiesis and Its Impact on Human Health. Annu. Rev. Med. 74, 249–260 (2023).

18. Jaiswal, S. & Ebert, B. L. Clonal hematopoiesis in human aging and disease. Science 366, eaan4673 (2019).

19. Jaiswal, S. et al. Age-Related Clonal Hematopoiesis Associated with Adverse Outcomes. N Engl J Med 371, 2488–2498 (2014).

20. Bick, A. G. et al. Inherited causes of clonal haematopoiesis in 97,691 whole genomes. Nature 586, 763–768 (2020).

21. Jaiswal, S. Clonal hematopoiesis and non-hematologic disorders. Blood blood. 2019000989 (2020) doi:10.1182/blood.2019000989.

22. Zhao, K. et al. Somatic and Germline Variants and Coronary Heart Disease in a Chinese Population. JAMA Cardiol 9, 233 (2024).

23. Zhao, K. et al. Clonal Hematopoiesis and Risk of Stroke: Evidence From Over 800 000 Individuals Across 3 Cohorts. Stroke STROKEAHA. 125. 054128 (2026) doi:10.1161/STROKEAHA.125.054128.

24. Bowman, R. L., Busque, L. & Levine, R. L. Clonal Hematopoiesis and Evolution to Hematopoietic Malignancies. Cell Stem Cell 22, 157–170 (2018).

25. Natarajan, P. Genomic Aging, Clonal Hematopoiesis, and Cardiovascular Disease. ATVB 43, 3–14 (2023).

26. Koh, Y., Tengesdal, I. W. & Jaiswal, S. Clonal Hematopoiesis in Nonmalignant Disease: Functional Consequences of Mutated Immune Cells by Clonal Hematopoiesis in the Diseased Tissue.

27. Hajishengallis, G. & Chavakis, T. Inflammageing and clonal haematopoiesis interplay and their impact on human disease. Nat Rev Mol Cell Biol 27, 377–393 (2026).

28. Bowman, G. I., Seymour, N., Reynolds, C. & Evans, L. M. Development and validation of an electronic health record-based frailty index in the UK Biobank. The Journals of Gerontology: Series A 81, glag131 (2026).

29. Street, A., Maynou, L., Blodgett, J. M. & Conroy, S. Association between Hospital Frailty Risk Score and length of hospital stay, hospital mortality, and hospital costs for all adults in England: a nationally representative, retrospective, observational cohort study. The Lancet Healthy Longevity 6, 100740 (2025).

30. Mak, J. K. L. et al. Large-scale genome-wide analyses with proteomics integration reveal novel loci and biological insights into frailty. Nat Aging 5, 1589–1600 (2025).

31. Walsh, B. et al. Frailty transitions and prevalence in an ageing population: longitudinal analysis of primary care data from an open cohort of adults aged 50 and over in England, 2006–2017. Age and Ageing 52, afad058 (2023).

32. Sekar, A. et al. Mosaic chromosomal alterations (mCAs) in individuals with monoclonal B-cell lymphocytosis (MBL). Blood Cancer J. 14, 193 (2024).

33. Kuo, C.-L., et al. A proteomic signature of healthspan. Proc. Natl. Acad. Sci. U.S.A. 122, e2414086122 (2025).

34. Pershad, Y. et al. Correlates and consequences of clonal hematopoiesis expansion rate: a 16-year longitudinal study of 6976 women. Blood 146, 1078–1087 (2025).

35. Immune Response to High-Dose vs. Standard Dose Influenza Vaccine. https://reporter.nih.gov/project-details/8891347 (2015).

36. Whitehead, J. C. et al. A Clinical Frailty Index in Aging Mice: Comparisons With Frailty Index Data in Humans. The Journals of Gerontology: Series A 69, 621–632 (2014).

37. McClatchy, J. et al. Clonal hematopoiesis related TET2 loss-of-function impedes IL1β-mediated epigenetic reprogramming in hematopoietic stem and progenitor cells. Nat Commun 14, 8102 (2023).

38. Genovese, G. et al. Clonal Hematopoiesis and Blood-Cancer Risk Inferred from Blood DNA Sequence. N Engl J Med 371, 2477–2487 (2014).

39. Arends, C. M. et al. Associations of clonal hematopoiesis with recurrent vascular events and death in patients with incident ischemic stroke. Blood 141, 787–799 (2023).

40. Ahn, H.-J. et al. Clonal haematopoiesis of indeterminate potential and atrial fibrillation: an east Asian cohort study. Eur Heart J 45, 778–790 (2024).

41. Schuermans, A. et al. Clonal haematopoiesis of indeterminate potential predicts incident cardiac arrhythmias. European Heart Journal 45, 791–805 (2024).

42. Shyr, D. et al. Clonal Hematopoiesis and Cardiovascular Disease Risk After Cancer Therapy in Patients With Solid Tumors.

43. Kestenbaum, B. et al. Clonal Hematopoiesis of Indeterminate Potential and Kidney Function Decline in the General Population. American Journal of Kidney Diseases 81, 329–335 (2023).

44. Vlasschaert, C. et al. Clonal hematopoiesis of indeterminate potential is associated with acute kidney injury. Nat Med 30, 810–817 (2024).

45. Wong, W. J. et al. Clonal hematopoiesis and risk of chronic liver disease. Preprint at 10.1101/2022.01.17.22269409 (2022).

46. Kim, P. G., et al. *Dnmt3a* -mutated clonal hematopoiesis promotes osteoporosis. Journal of Experimental Medicine 218, e20211872 (2021).

47. Hoogendijk, E. O. et al. Frailty: implications for clinical practice and public health. The Lancet 394, 1365–1375 (2019).

48. Ijaz, N. et al. Interventions for Frailty Among Older Adults With Cardiovascular Disease. Journal of the American College of Cardiology 79, 482–503 (2022).

49. Zekavat, S. M. et al. Hematopoietic mosaic chromosomal alterations increase the risk for diverse types of infection. Nat Med 27, 1012–1024 (2021).

50. Lin, S.-H. et al. Incident disease associations with mosaic chromosomal alterations on autosomes, X and Y chromosomes: insights from a phenome-wide association study in the UK Biobank. Cell Biosci 11, 143 (2021).

51. Sano, S. et al. Hematopoietic loss of Y chromosome leads to cardiac fibrosis and heart failure mortality. Science 377, 292–297 (2022).

52. Pershad, Y. et al. Mutation-specific impairment of TET2 and DNMT3A enzymatic activity predicts clonal hematopoiesis disease risk.

53. Carter, P. R. et al. Common clonal hematopoiesis driver mutations have disparate effects on macrophage cytokines, clonal expansion, and atherogenesis.

54. Belizaire, R., Wong, W. J., Robinette, M. L. & Ebert, B. L. Clonal haematopoiesis and dysregulation of the immune system. Nat Rev Immunol 23, 595–610 (2023).

55. Robertson, N. A. et al. Longitudinal dynamics of clonal hematopoiesis identifies gene-specific fitness effects. Nat Med 28, 1439–1446 (2022).

56. Qiu, R., Zhang, D. & Feng, H. Immunosenescence as a driver of the transition from frailty to multimorbidity. Front. Immunol. 17, 1810241 (2026).

57. Caiado, F. et al. Aging drives *Tet2 +/−* clonal hematopoiesis via IL-1 signaling. Blood 141, 886–903 (2023).

58. Jaiswal, S. & Libby, P. Clonal haematopoiesis: connecting ageing and inflammation in cardiovascular disease. Nat Rev Cardiol 17, 137–144 (2020).

59. Ramirez, A. H. et al. The All of Us Research Program: Data quality, utility, and diversity. Patterns 3, 100570 (2022).

60. The All of Us Research Program Genomics Investigators et al. Genomic data in the All of Us Research Program. Nature 627, 340–346 (2024).

61. The UK Biobank Whole-Genome Sequencing Consortium et al. Whole-genome sequencing of 490,640 UK Biobank participants. Nature https://doi.org/10.1038/s41586-025-09272-9 (2025) doi:10.1038/s41586-025-09272-9.

62. Pendergrass, S., Dudek, S. M., Roden, D. M., Crawford, D. C. & Ritchie, M. D. Visual integration of results from a large DNA biobank (BioVU) using synthesis-view. Biocomputing 2011 265–275 (2010) doi:10.1142/9789814335058_0028.

63. Vlasschaert, C. et al. A practical approach to curate clonal hematopoiesis of indeterminate potential in human genetic data sets. Blood 141, 2214–2223 (2023).

64. Zhao, K. et al. Interleukin-17 receptor-A signalling: atheroprotective role in JAK2 clonal haematopoiesis. European Heart Journal ehaf 737 (2025) doi:10.1093/eurheartj/ehaf737.

65. Jaiswal, S. et al. Clonal Hematopoiesis and Risk of Atherosclerotic Cardiovascular Disease. N Engl J Med 377, 111–121 (2017).

66. Loh, P.-R., Genovese, G. & McCarroll, S. A. Monogenic and polygenic inheritance become instruments for clonal selection. Nature 584, 136–141 (2020).

67. Zhao, K. et al. Genetic drivers and clinical consequences of mosaic chromosomal alterations in 1 million individuals. Preprint at 10.1101/2025.03.05.25323443 (2025).

68. Zaslavsky, O. et al. Comparison of Frailty Phenotypes for Prediction of Mortality, Incident Falls, and Hip Fracture in Older Women. J American Geriatrics Society 64, 1858–1862 (2016).

69. Pershad, Y. et al. Correlates and consequences of clonal hematopoiesis expansion rate: a 16-year longitudinal study of 6976 women. Blood 146, 1078–1087 (2025).

70. Vlasschaert, C. et al. Association of Clonal Hematopoiesis of Indeterminate Potential with Worse Kidney Function and Anemia in Two Cohorts of Patients with Advanced Chronic Kidney Disease. JASN 33, 985–995 (2022).

71. Rockwood, K. & Theou, O. Using the Clinical Frailty Scale in Allocating Scarce Health Care Resources. Can Geri J 23, 254–259 (2020).

